# A novel age-informative polygenic score improves predictive ability for kidney function and kidney function decline

**DOI:** 10.64898/2026.01.22.26344420

**Authors:** Janina M. Herold, Simon Wiegrebe, Barbara Thorand, Thomas W. Winkler, Christian Gieger, Florian Hartig, Merle Behr, Annette Peters, Helmut Küchenhoff, Iris M. Heid

**Author notes:** Corresponding Author: Prof. Dr. Iris M. Heid, Department of Genetic Epidemiology, University of Regensburg, Franz-Josef-Strauss-Allee 11, 93053 Regensburg, Germany, Tel.: +49 (0)941/944-5210.

## Abstract

Polygenic scores (PGSs) are widely used to summarize the joint genetic effects for disease-related traits. However, while age-dependent genetic effects are increasingly recognized, their integration into PGSs remains underexplored. Kidney function, assessed by estimated glomerular filtration rate (eGFR), has strong age-related genetic effects, and prediction of kidney function decline is an unmet need.

We develop an age-informative PGS for quantitative traits by generating age-specific weights via main and interaction effects and compare its performance to the age-agnostic PGS in theory and real data of eGFR. We test PGSs across 282 kidney function SNPs in cross-sectional and longitudinal data from UK Biobank (n=348,275, m=1,520,382) and independent population-based individuals aged 25 to 98 years (KORA&AugUR; n=9,057, m=16,804).

In theory and real data, we illustrate that ignoring age mis-specifies genetic effects. The age-informative PGS has better performance than the age-agnostic PGS in young and old individuals (KORA&AugUR: 6.3% versus 5.9% of eGFR variance in 25-to 45-year-old, 2.3% versus 1.8% in 75-to 98-year-old). The PGS based on interaction effects explains more of the eGFR-decline variability than the age-agnostic PGS. The highest versus lowest PGS quintile predict eGFR-decline of −0.88 (95%-CI=[−0.93;−0.83]) versus −0.75 ml/min/1.73m^2^ (95%-CI=[−0.79;−0.71]) on the population-level, similar to strata by acquired risks like diabetes, obesity or albuminuria. Prediction of eGFR-decline on the individual level remains challenging by both, genetic or acquired risks.

Overall, we provide a simple approach to an age-informative PGS for quantitative disease traits and illustrate its chances and challenges for predicting kidney function and kidney function decline.

## Introduction

Polygenic scores (PGSs) are a powerful means to increase the predictive ability of non-genetic models for disease traits^1, 2^. They facilitate risk stratification at the population level and show promise as clinical tools for estimating an individual’s susceptibility to specific traits and related health risks^1^. A conventional PGS uses GWAS-identified genetic variants associated with a disease or quantitative disease trait. The individual’s PGS reflects the person’s genetic profile for the disease or trait^3^.

In recent years, more and more GWAS consider gene-environment interaction (GxE), reflecting complex genetic mechanisms underlying disease traits^4^. Accounting for such interactions in PGSs can be expected to improve the predictive ability for such traits^5^. Most currently available approaches to implement such interactions in PGSs focused on binary disease outcomes or binary environmental factors. For genetic interactions with a binary time-independent factor such as sex, one way to account for interaction effects in PGSs is to calculate sex-specific PGSs. For blood pressure traits, for instance, sex-specific PGSs have been shown to improve predictive power for both women and men^6^. For genetic interactions with a quantitative environmental factor, most previous work focused on the (interaction) effects of genetics and age on disease, e.g., on prostate cancer^7^ or Alzheimer’s disease^8^. The genetic effects on these diseases have been found to decrease by age due to various other lifetime risks that increase over age. Recent work proposed an approach to properly integrate GxE effects in PGS and its impact on statistical power and type I error when testing the PGS effect on binary as well as quantitative disease trait^5^. This approach requires the generation of two PGSs, one for the main effect and one for the interaction effect and was exemplified on traits like body-mass-index or low-density lipoprotein and genetic interaction with lifestyle factors. The application of such GxE informative PGSs may have implications on personalized medicine but warrants an evaluation of their performance when compared to age-agnostic PGS.

An important application where the combination of genetic profile and GxE analyses may be key to improve prediction is the estimated glomerular filtration rate (eGFR), which is a marker of kidney function^9^. Kidney function naturally declines with age due to loss of nephrons^10^, and accelerated decline can lead to kidney failure necessitating dialysis or kidney transplantation. Prediction of kidney function and kidney function decline is clinically highly relevant as the current means to predict a person’s kidney function or its decline are very limited^11,10,12,13^. Hundreds of independent genetic variants have been identified to be associated with eGFR in cross-sectional data GWAS of about 1 million individuals^14, 15^. A recent study using longitudinal data with 1.5 million eGFR values from UK Biobank identified genetic effects on eGFR that strongly depended on age and all these genetic effects increased with age^16^. An age-informative PGS could improve the prediction of kidney function and kidney function decline compared to a conventional age-agnostic PGS. However, it is unclear how exactly to generate an age-informative PGS in this setting and how much this would improve predictive ability.

We thus aimed to develop an approach that integrated age effects into a PGS for quantitative traits and to derived age-informed genetic prediction of kidney function and kidney function decline. Specifically, we (i) provide a concept for a novel age-informative PGS for quantitative traits and (ii) illustrate its impact in a simplified theoretical case study. (iii) We compare the age-informed PGS with the age-agnostic PGS as well as a previously suggested GxE approach regarding predictive ability for kidney function and kidney function decline. For this, we use longitudinal and cross-sectional data from UK Biobank (40-78 years, n~300,000) and KORA&AugUR (25-98 years, n~9,000). (iv) Finally, we determine the ability of the age-informative PGS to predict kidney function and kidney function decline on the population- and individual-level.

## Methods

### General concept of an age-agnostic and an age-informative PGS for quantitative traits including the novel approach

A PGS of an individual counts the number of “bad” alleles across a set of SNPs, where the “bad” allele has been shown to be associated with a trait into the disease-related direction. On the example of kidney function, the “bad” allele is associated with lower kidney function. A conventional PGS across *k* independent SNPs is generated as 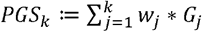, where *G*_*j*_ denotes the individual’s dosage of the bad allele for the *j*^*th*^ SNP (*j*=1, …, *k*). Weights are derived from the bad alleles’ association estimates, 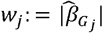, from linear regression GWAS in cross-sectional data,

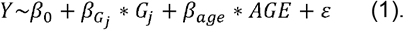

Age-dependent genetic effects can be estimated via linear regression with interaction term,

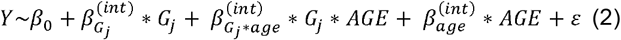

Where 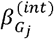 is the main genetic effect and 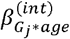 is the GxAge interaction effect (i.e. change of genetic effect per allele and year of age). When the variable AGE is centered at, e.g., 50 years (denoted as AGE50 in the following), the main effect is the effect of *G*_*j*_ for 50-year-old individuals. Similarly, age-dependent effects can be derived as GxAge interaction from longitudinal data via linear mixed models (LMMs) on a time-dependent Y_t_ with random intercepts (RI) and random slopes (RS) (e.g.^16^). While the direction of the marginal genetic effect estimates, 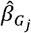, can be aligned to the “bad” allele, the directions of 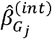 and 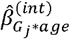 from (2) need to be evaluated in the specific context. Age-specific genetic effects can be derived as

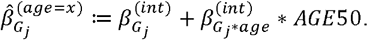

The marginal effect reflects the age-specific genetic effects averaged across all ages of the individuals in the dataset. When 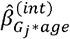 is not significantly different from 0, the marginal effect is the same (except for statistical error) as the main effect and age-specific effects at any age.

We postulate that an age-informative PGS, which quantifies a person’s genetic profile to predict Y at a certain age, can be generated by age-specific weights, 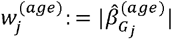, as 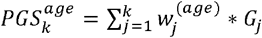. Age-specific weights can also be separated into weights based on main effects, 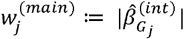, and weights based on interaction effects, 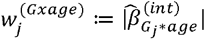. These can be used to generate a “main-effect PGS”, 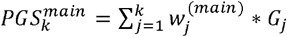 and an “interaction-effect PGS”, 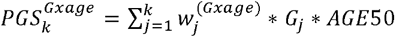.

The unit of a PGS is not interpretable when weights are beta-estimates (“unscaled”), e.g.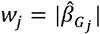. To yield interpretable units, we divide these weights by their average across SNPs, e.g. divide by 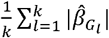, to yield “scaled weights”. Then, one PGS unit reflects one allele of “average effect”. In the following, we refer to scaled weights, if not noted otherwise.

### A simplified case study to illustrate differences in genetic effects and PGS performance without and with accounting for age

To derive an expectation for the impact of accounting for age, we generated a simplified case study: we assumed five SNPs without interaction on trait 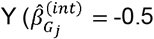 trait units per allele, 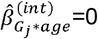 trait units per allele and year) and five SNPs with age interaction (zero effect until age 50 years, 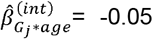 trait units per allele and year afterwards. In a thought experiment, we considered an equal number of persons of aged 40, 50, … or 80 years and derived age-specific effects and marginal effects across ages. Using these effects divided by their average as (scaled) weights, we derived the age-informative and age-agnostic PGSs across the 10 SNPs, *PGS*_10_ or 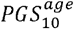, and considered them “tested” via linear regression in individuals of the specific age (*x* years). We used the average of the weights of the 10 SNPs, *w*_*j*_ or 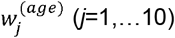 as proxy for the per unit effect of the age-agnostic or age-informative PGS, respectively. We assumed equal variance of SNPs (*var*(*Gj) = var* (*G*), *j*=1, …, *k, k*=10) and constant trait variance over age, given that SNP effects on Y were small. The variance of the age-agnostic or age-informative PGS (at age *x* years) can then be derived as

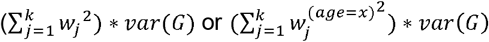

and the trait variance explained by the PGS, R^2^(PGS), as

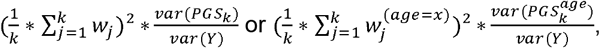

respectively, which is a measure of predictive ability.

### UK Biobank and KORA&AugUR with cross-sectional and longitudinal data on kidney function and genetics

We used two datasets to evaluate PGS performance for kidney function: UK Biobank and KORA&AugUR. UK Biobank included 500,000 participants aged 40-69 years at baseline study center visit (mean=57 years). We derived longitudinal data on eGFR combining serum creatinine measurements from study-center visits (Beckman Coulter assay) with electronic health records (eHRs) from general practitioners (“GP-clinical”, UK Biobank application number 20272) as described previously^17^. KORA&AugUR is a combined dataset from three German population-based cohort studies^18, 19^ with longitudinal data on eGFR as established previously^20^. The joint dataset covers an age range of 25 to 95 years at baseline (mean=60 years) with up to four visits. Serum creatinine concentrations were measured by enzymatic assays or modified Jaffé^20^.

Inclusion/exclusion criteria were similar in both datasets: we included unrelated, European-ancestry individuals and excluded eGFR values (i) at and after recorded dialysis, (ii) <6 months before and all values after kidney transplantation or nephrectomy, (iii) after an observed eGFR value <15 mL/min/1.73□m^2^. In addition, in UK Biobank, values were excluded +/-6 months of recorded acute kidney injury and +/-10 months of recorded pregnancy. Information on dialysis, kidney transplantation, and nephrectomy was derived from self-reports (KORA&AugUR) or medical records (GP-CTV3 and read-V2 codes, UK Biobank^17^). We derived eGFR based on creatinine (CKD-EPI 2021^21^) and winsorized values <15 and >200□mL/min/1.73□m^2^. Age at the time of blood sampling was used as the person’s age.

These data enabled longitudinal analyses (UK Biobank, n=348,275, m=1,520,382; KORA&AugUR, n=9,057, m=16,804) and cross-sectional analyses. For cross-sectional analyses over all ages, we used the data from baseline study center visit. For cross-sectional analyses stratified by age groups (25-45 “young”; 45-65 “middle-aged”, 65-78 years; “old-aged”; additionally 75-98 years, “very old-aged”, in KORA&AugUR), we enriched by longitudinal eGFR values for young and old individuals (eHR-based in UK Biobank; study center follow-up visits in KORA&AugUR): we used the first eGFR-value for a person between 25 and 45 years or the last value between 65 and 78 years; for the middle-aged group, we used the value assessed closest to 60 years. This ensured no more than 1 value per individual and group but allowed individuals to appear in more than one group.

For both datasets, we had individual participant data with genome-wide SNP information. UK Biobank data was imputed via HRC and UK10K reference panels^22,23^ as described in ^16^; KORA&AugUR was imputed via HRC panel^24, 20, 25^. We selected 595 SNPs previously identified for genome-wide significant association with creatinine-based eGFR (P<5×10-8; cross-sectional, CKDGen and UK Biobank^26^; n=1,201,929). These SNPs were rather common and well-imputed in UK Biobank and KORA&AugUR (MAF>1%, INFO-score≥ 0.6); 12 of these SNPs had significant interaction with age for eGFR in UK Biobank (P_Gj*age_ <0.05/595). We restricted further to 282 SNP associated with cystatin-based eGFR (P<1×10^−5^, cross-sectional UK Biobank^27^) to ascertain SNPs as kidney function relevant.

### UK Biobank data to evaluate genetic effects for kidney function without and with accounting for age

For the 282 kidney function SNPs, we used marginal (baseline study center, linear regression, n=254,068), main and GxAge interaction effects (longitudinal data, LMM with RI and RS; n=348,275; m=1,520,382) from UK Biobank as published previously^16^. Effect alleles were set to the eGFR-lowering allele of the marginal effect (i.e. 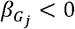). We compared marginal effects with age-specific effects on eGFR derived as 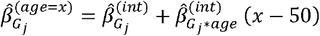 for *x* = 40, 50, 60, 70, or 80 years. We reviewed positive age-specific effects 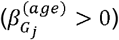 regarding the age where the “bad” allele switched (i.e. *x* where 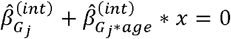).

Since a PGS weighs in the genetic effects relative to each other, the correlation in the effects used as weights (not their absolute value) drives the differences in PGSs. The scaling of weights does not affect the correlation. We thus compared the 282 age-agnostic genetic effects with age-specific effects and interaction effects using Spearman correlation.

### Calculating four types of PGSs for kidney function and deriving predictive ability in UK Biobank and KORA&AugUR

Using individual participant data from UK Biobank and KORA&AugUR for the 282 kidney function SNPs, we generated four types of PGSs: (i) the age-agnostic PGS, *PGS*_282_ using marginal effects for weights, (ii) our age-informative PGS, 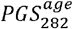, using age-specific genetic effects, (iii) a main-effects PGS, 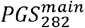, and (iv) an interaction-effects PGS, 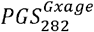, (**Figure 1**). We tested the PGSs and derived their predictive ability in UK Biobank (weight-identifying dataset) and in KORA&AugUR (independent data).

**Figure 1.**
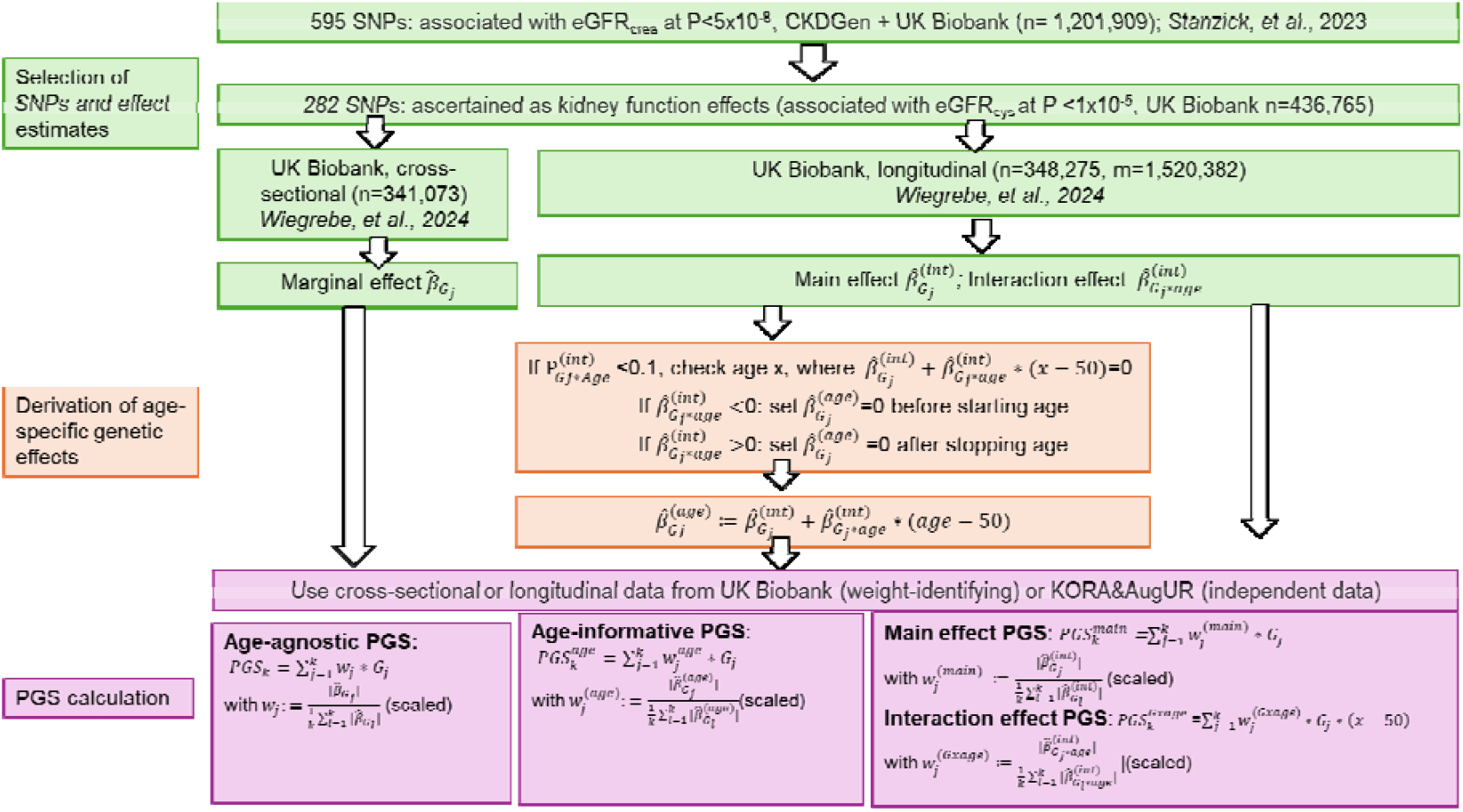
Workflow of generating four types of PGSs across 282 SNPs associated with kidney function. Based on 595 SNPs identified for genome-wide significant association with eGFR_crea_ (*Stanzick et al*., *2023*), we defined a subset of 282 kidney function SNPs ascertained for association with eGFR_cys_ in UK Biobank. Published summary statistics provided marginal effect estimates ( from cross-sectional analysis and main () and interaction effects ( from longitudinal analysis in UK Biobank (*Wiegrebe et al*., *2024)*. For SNPs with observed interaction with age, we derived a starting age when or a stopping age when and set to 0 before starting age or after the stopping age, respectively. Based on main and interaction effects, we derived age-specific effect estimates (. In UK Biobank and KORA&AugUR data, we generated an age-agnostic PGS based on, an age-informative PGS based on, and a main-effect PGS and interaction-effect PGS based on or, respectively.

First, in cross-sectional data, we applied linear regression for the age-agnostic PGS,

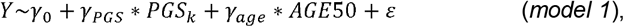

(R function *lm()*) and for the age-informative PGS,

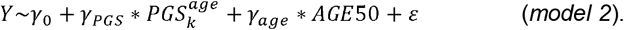

We derived the models’ total R^2^, which provides the proportion of trait variance explained by the PGS and AGE50. We compared these with the R^2^ from a reduced model (only AGE50),

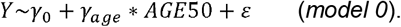

We also derived partial R^2^ for the PGS, which provides the proportion of explained variance attributable to the PGS (on top of AGE50). For this, we subtracted the R^2^ of the reduced model (*model 0*) from the total R^2^ as

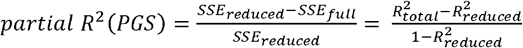

where SSE denotes the sum of squared deviations between observed (*Y*) and predicted values (*Ŷ*) in the reduced (*SSE*_*reduced*_) or the full model (*SSE*_*full*_).

Second, again using cross-sectional data, we compared the predictive ability of our age-informative PGS with the joint predictive ability of two separate PGSs, main-effect PGS and interaction-effect PGS, in a model according to *Jayasinghe et al*.^5^,

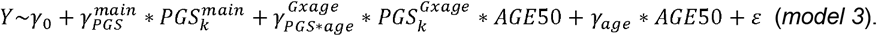

Since the sums in

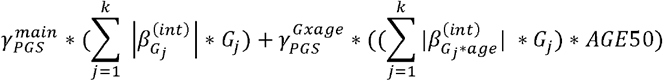

can be re-arranged to

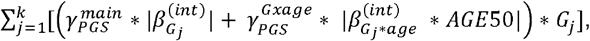

*model 3* is similar as *model 2*, except it allows for separate *γ*-estimates for main and interaction effects (**Supplemental Note 1**). Furthermore, we added 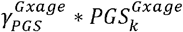 to *model 3* following the recommendation from *Jayasinghe et al*.^5^ (*model 4*). We derived total R^2^ and partial R^2^ for the joint effect of 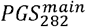 and 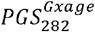 by subtracting the R^2^ of the reduced model (*model 0*) as described above.

Third, we tested the performance of PGSs in longitudinal data using LMMs with RI and RS. For the age-informative PGS, we applied a LMM corresponding to *model 2*,

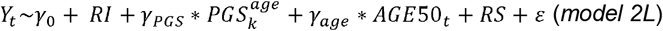

(R function *lmer()*, lme4 package) and, for the age-agnostic PGS, the LMM analogous to *model 1* (*model 1L*). We also extended *models 1L* and *2L* by adding the PGS interaction with age. We also tested the main effect and interaction effect PGSs in the models from *Jayasinghe et al*. ^5^,

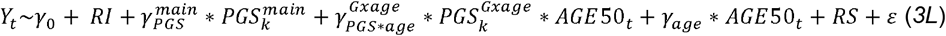

and adding 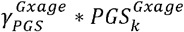 (*model 4L*). We derived marginal R^2^ for these LMMs, which reflected the proportion of trait variance explained by the fixed effects of all covariates, and partial R^2^ for joint effect of the PGSs by subtracting the marginal R^2^ from *Y*_*t*_~*AGE*50_*t*_ (reduced model) from the marginal R^2^ of the full model as described above.

### Comparing the interaction-effect PGS versus the age-agnostic PGS to predict kidney function decline

Given the clinical relevance of kidney function decline, we were particularly interested in the association and predictive ability of the PGS on eGFR-decline, as little is known about the joint genetic impact on eGFR-decline and how to generate a PGS for a trait change in general).

The study of risk factor association with eGFR-decline is typically conducted in longitudinal data using LMMs on time-dependent eGFR with RI and RS. Annual eGFR-decline is the change of eGFR by year of age in observational data when baseline is a random timepoint and does not mark an intervention; the interaction of a risk factor (here PGS) with age provides the risk factor association with eGFR-decline^16^. Thus, we considered the interaction-effect PGS, 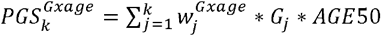, as appropriate PGS for eGFR-decline. The *Jayasinghe et al. model 3L* is then a straight-forward approach to quantify the PGS association with eGFR-decline by the interaction of 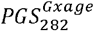 with age. This can be compared to the interaction of a conventional PGS (using marginal effects, *PGS*_282_) with age in the *extended model 1L*. However, these models involve interaction terms of PGS with age and thus do not provide interpretable partial R^2^ values for the PGS on eGFR-decline.

An alternative to study risk factor association with eGFR-decline is a two-step approach: (i) estimate person-specific slopes via “best linear unbiased predictors” (BLUPs)^28^ in longitudinal data using a LMM with RI and RS, *Y*_*t*_ ~*AGE*50_*t*_ and (ii) test the association of the PGS (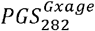 or 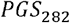) with person-specific slopes (PSS) via linear regression^29^,

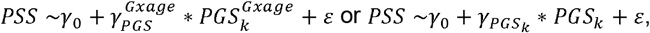

respectively. While this approach involves a bias towards the null in genetic effect estimates and leaves the uncertainty from step (i) unaccounted for in step (ii) [16], we apply this nevertheless to derive the explained variance (R^2^) of these univariable models as predictive ability of the PGSs on eGFR-decline.

### Prediction of eGFR and eGFR-decline on population- and individual-level based on PGS risk strata and other risk factors

For personalized medicine, it is important to understand whether genetic profiles can differentiate the prediction of eGFR values or eGFR-decline. Linear regression models (cross-sectional data, for eGFR) or LMM (longitudinal data, for eGFR-decline) can be used to derive mean levels of eGFR or eGFR-decline by genetic profile. For this, we analyzed KORA&AugUR, since UK Biobank was the weight-identifying dataset, cross-sectionally (n=254,068) to predict eGFR and longitudinally to predict eGFR-decline (n=348,275, m=1,520,382).

First, we compared extreme and mean genetic profiles of 0, 200, 285, 364, or 594 “bad” alleles (min possible, ~min observed, mean observed, ~max observed, or max possible, respectively). To predict eGFR for individuals at a certain age (*x* years) by genetic profile, we used results of the age-informative PGS in *model 2* and derived mean eGFR by PGS profile 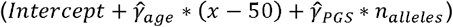. To predict annual eGFR-decline, we used results of the interaction effect PGS in *model 3L* and derived mean eGFR-decline by PGS profile 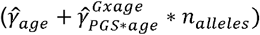.

Second, we stratified groups Q1-Q5 using PGS quintiles. For the age-informative PGS, we defined a quintile indicator function, 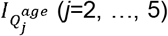, which takes on the value 1 if the individual’s age-informative PGS is in the *j*^th^ quintile. To predict eGFR, we then applied linear regression using these quintile indicators as covariates in cross-sectional data,

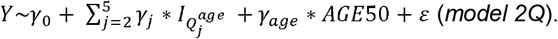

We derived mean eGFR for individuals at certain age (*x* years) in the low- or high-risk group (Q1 or Q5) as 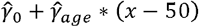 or 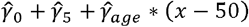, respectively. To predict eGFR-decline by quintiles of the interaction effect PGS, we defined 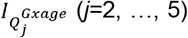 analogously and applied a LMM with RIs and RSs in longitudinal data,

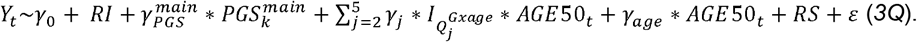

We derived mean eGFR-decline for Q1 or Q5 as 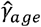 or 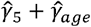, respectively.

Differences in mean values when comparing unfavorable versus favorable profiles reflect the impact on the population mean when the unfavorable profile could be modified to a favorable profile (prediction on population-level). We also derive corresponding 95%-CIs that account for the statistical uncertainty and 95%-prediction intervals that account additionally for the person variability (i.e. residual variance, for eGFR, or random slopes variability, for eGFR-decline). The 95%-prediction intervals by genetic risk group reflect the range of values that can be expected for 95% of individuals in the group (prediction on person-level; R function *predict()*).

Finally, to put predicted eGFR-decline for high- and low-genetic risk into perspective, we extended *model 3Q* by adding “non-genetic” risk factors (diabetes, albuminuria, and obesity) as well as their interaction with age into the model (*extended model 3Q*). We derived mean eGFR-decline, 95%-CIs, and 95%-prediction intervals by non-genetic and genetic risk groups using the effect of age and risk factor interaction with age. Diabetes was defined via self-report, antidiabetic medication intake (Anatomical Therapeutic Chemical classification ^30^), or HbA1c ≥6.5%. Albuminuria was derived from urinary albumin to urinary creatinine ratio (≥30 mg/g). Body-mass index (BMI), computed as measured weight (from each visit) divided by squared height (baseline visit), was used to define “overweight” (BMI ≥25 and <30 kg/m^2^) or “obesity” (≥30kg/m^2^). Diabetes, albuminuria, overweight and obesity status assessed at the timepoint of the eGFR assessment was used for analyses.

## Results

### A simplified case study illustrates mis-specified genetic effects and lower predictive ability of the PGS when ignoring age

To demonstrate how genetic effects on a trait are mis-specified and predictive ability compromised when ignoring age-dependent effects, we used a simplified case study.

First, we considered a theoretical SNP simulating an age-dependent effect on a trait Y (zero effect until age of 50 years, −0.05 units per allele and year afterwards). The marginal effect of −0.6 trait units per allele deviated substantially from age-specific effects in young and old individuals (e.g. 0 or −1.5 trait units per allele in 40- or 80-year-old, respectively; **Figure 2a**).

**Figure 2.**
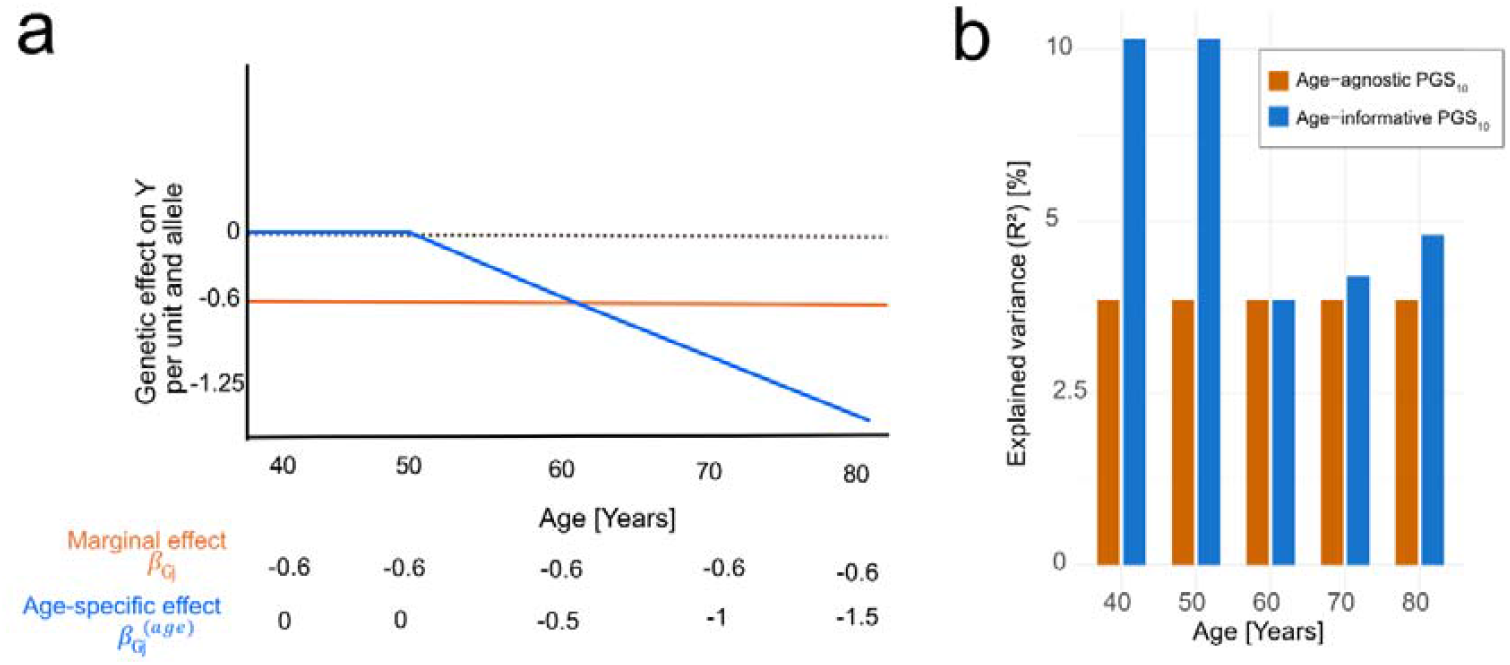
Misspecified genetic effects on a quantitative trait when ignoring age-dependency and its impact on prediction by PGSs in a theoretical example. First, we considered a theoretical SNP with an age-dependent effect on Y with zero effect until 50 years and then 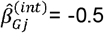 trait units per allele and year. Shown is (**a**) the age-specific effect estimates for this SNP 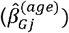 for a 40-, 50-, 60-, 70-, or 80-year-old person (blue) contrasted to the marginal effect assuming a sample with equal number of individuals of each of these ages 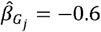 units per allele, orange). Second, we considered five such SNPs and five SNPs with age-independent effects (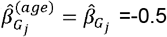 trait units per allele). We generated age-agnostic and age-informative PGSs across these 10 SNPs. Shown is (**b**) the percentage of explained variance 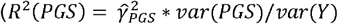 for the different ages when using an age-agnostic PGS (orange) or an age-informative PGS (blue). Variance of *G*_*j*_ is assumed to be constant across SNPs and variance of Y to be constant across ages. The average effect across the 10 SNPs reflects the per unit effect of the (scaled) PGS 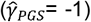. Detailed values are provided in **Supplemental Table S1**.

Second, we considered 5 such SNPs and 5 SNPs with age-independent effects (−0.5 trait units per allele). The age-agnostic PGS effect of −0.5 trait units per average allele deviated substantially from the age-informative PGS effects in young and old individuals (−0.25 or −1 trait units per average allele for 40- or 80-year-old, respectively; **Supplemental Table S1**). The R^2^ was 3.8% for individuals at any age for the age-agnostic PGS and 7.7% or 5.2% for 40- or 80-year-old individuals, respectively, for the age-informative PGS (**Figure 2b; Supplemental Table S1**). This illustrated higher predictive ability of the age-informative instead of the age-agnostic PGS for younger and older individuals. This also demonstrated the age-agnostic PGS to perform well for individuals at mean age (60 years; R^2^=3.8%).

### Data from UK Biobank shows mis-specified genetic effects when ignoring age based on 282 SNPs associated with kidney function

We evaluated how genetic variant effects for eGFR were mis-specified when ignoring age-dependency for the 282 kidney function SNPs, deriving age-specific effects from main and interaction estimates and compared these to marginal effects ^16^.

The example of the 12 SNPs identified previously with GxAge interaction^16^ illustrated the near zero effects for individuals aged 40 years and then, for the effect allele (eGFR-lowering for marginal effect) accelerated decline by age (“decline effect SNPs”, **Figure 3a**). This also showed that some SNP effects intersected the zero-line at young age, which we considered a result of model misspecification by the linear GxAge relationship when genetic effects were in fact zero until a starting age (intersection with zero-line). The difference between marginal and age-specific effects illustrated overestimation of effects in young and underestimation in old individuals when age is ignored (**Figure 3b; Supplemental Table S2**). Age-specific effects for 60-year-old and marginal effects were the same (**Figure 3b**), in line with mean age in this data being 60.9 years (**Table 1**).

**Table 1.**
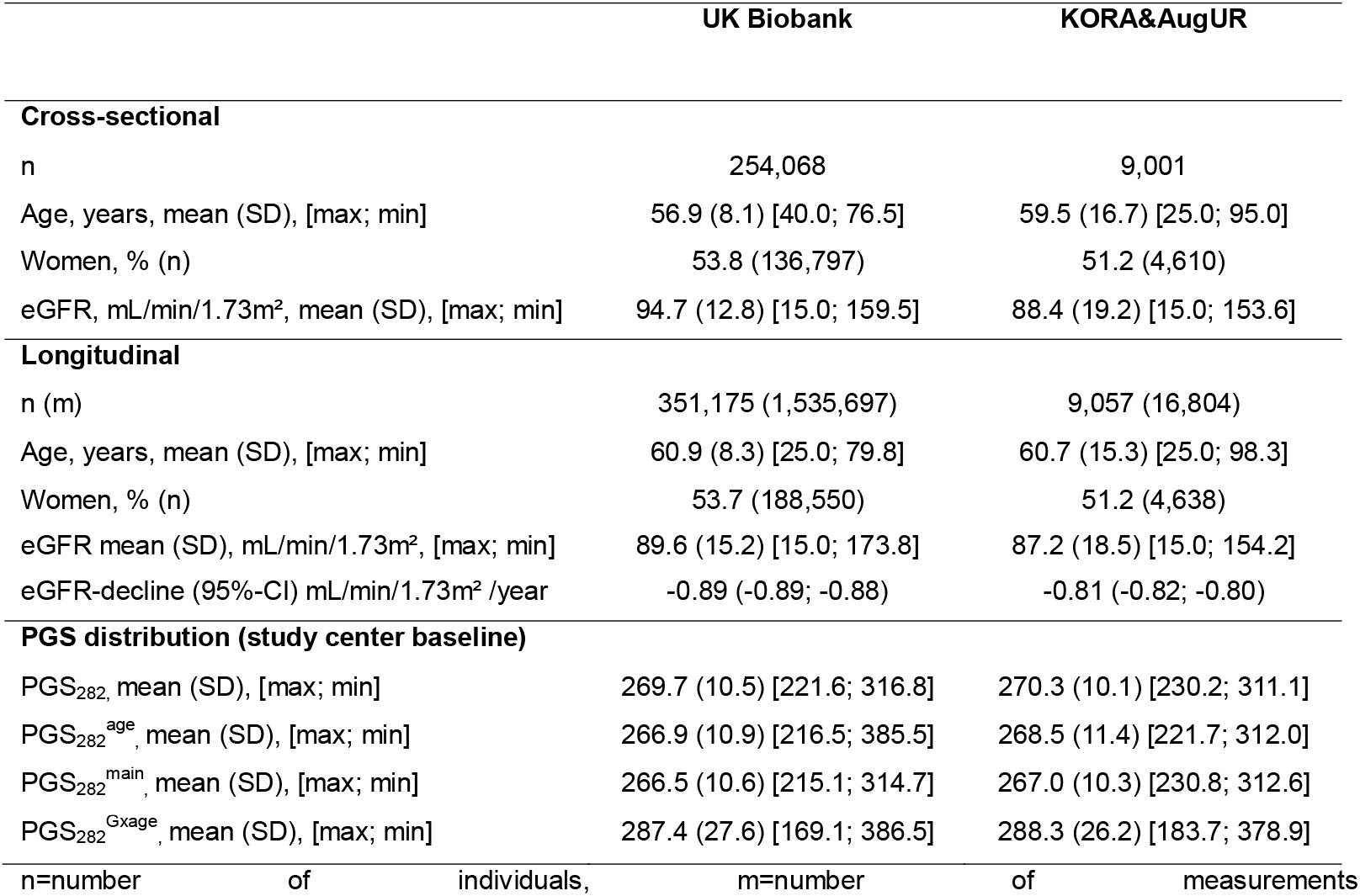
Participant characteristics of the analyzed samples. Shown are participant characteristics for UK Biobank and KORA&AugUR for cross-sectional and longitudinal analyses. Continuous variables are presented as mean with standard deviation (SD) in parentheses and minimum and maximum in brackets; categorical variables are shown as percentage and counts (n). Additionally, the estimated annual eGFR-decline resulting as an age effect from a linear mixed model in longitudinal data, *eGFR*_*t*_~*AGE*50 is provided with 95%-CIs in parentheses. We also show descriptive statistics of the four PGSs across the 282 kidney function SNPs (age-agnostic, PGS_282,_ age-informative, PGS_282_^age^, main effect PGS, PGS_282_^main^, and interaction effect PGS, PGS_282_^Gxage^.

**Figure 3.**
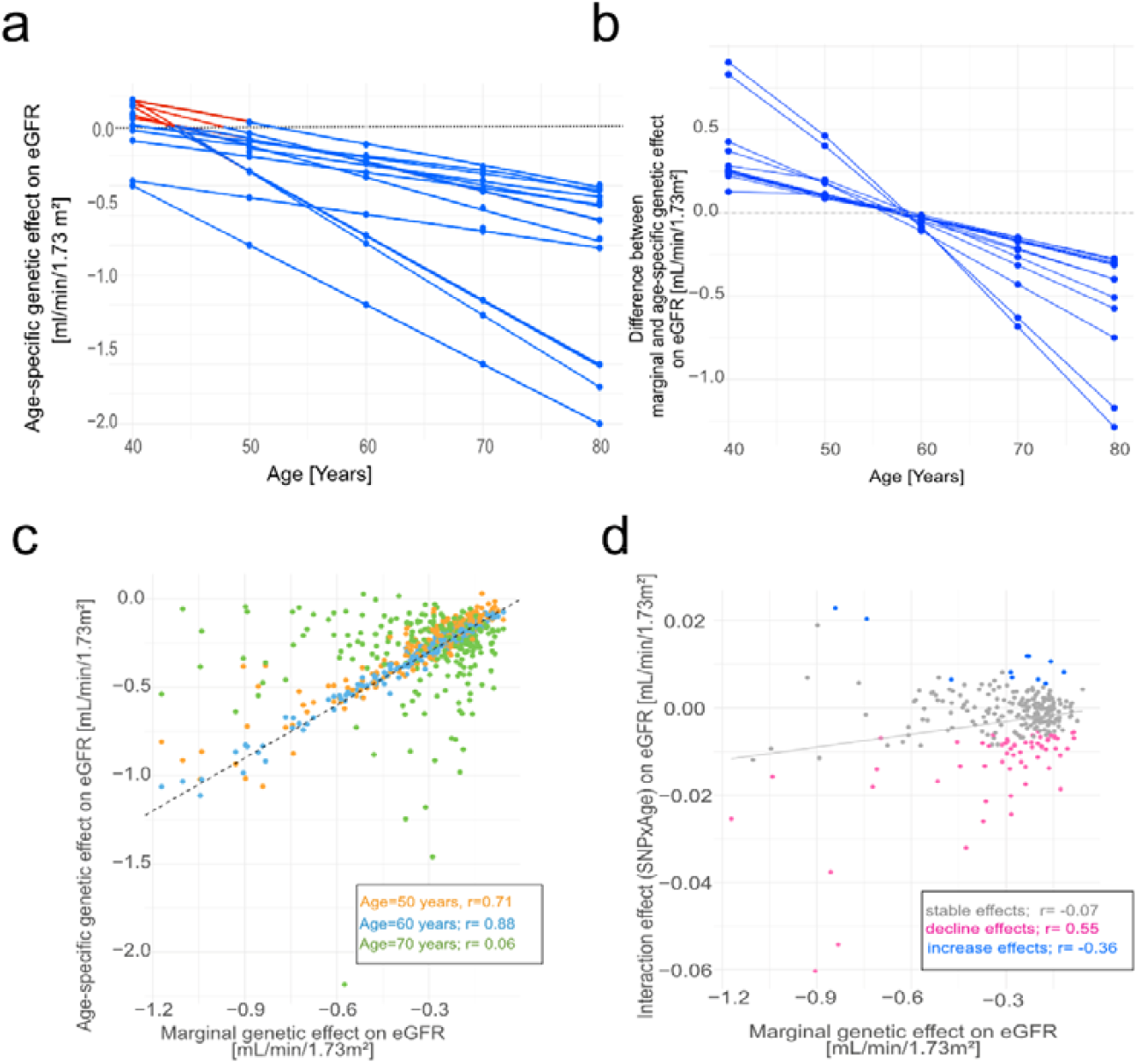
Misspecified genetic effects for eGFR when ignoring age-dependency in UK Biobank. Based on published beta-estimates (*Wiegrebe et al*., *2024)*, we derived age-specific genetic effects for the 282 kidney function SNPs as 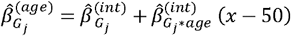 for x=40, 50, 60, 70, and 80 years of age. On the example of 12 SNPs known for interaction with age, we show (**a**) age-specific beta-estimates, 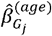, by age (blue points). We considered the intersection of the blue lines with the x-axis as age-of-onset of the genetic effects (red) and set 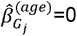 until this “starting age” (x where 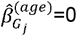). We also show (**b**) the difference between the marginal 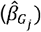 and the age-specific effect 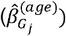 for the 12 SNPs. For all 282 SNPs, we contrast (**c**) age-specific effects, 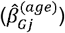 with marginal effects 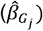. At age = 50 years, 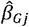 corresponds to the main effect 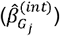 derived from a model with age centered at 50 years. We also contrast (**d**) interaction effects 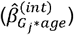 with marginal effects 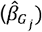 for the different types of SNP effects (stable: P_G*age_ ≥0.1; decline: 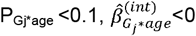; increase: 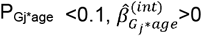. In (**c**) and (**d**), the correlation coefficient r refers to Spearman’s rank correlation.

Among the 282 SNPs, we observed (i) 63 SNPs with negative interaction 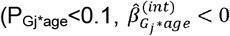 “decline effect SNPs”), (ii) 12 SNPs with positive interaction 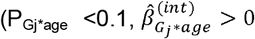 “increase effect SNPs”), and (iii) 208 SNPs without interaction (P_G*age_ ≥0.1; “stable effect SNPs”). We found positive effects, 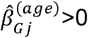, for 49 of 63 “decline” effect SNPs (intersections of zero-line between 25 and 48 years) and for 10 of the 11 “increase effect SNPs (intersections between 70 and 100 years; **Supplemental Data 1**). Since the intersections of the zero-line were found for “decline” effect SNPs at young age and for “increase” effect SNPs at old age, we considered these as the age where the SNP effect started or stopped, respectively (setting 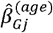 to zero until starting or after stopping age). The difference of marginal and age-specific effects across the 282 SNPs varied between 0.13 and 0.91 mL/min/1.73m^2^ per allele in 40-year-old and between −1.3 and −0.3 in 80-year-old individuals (**Supplemental Data 1**). This highlights the misspecification of SNP effects on eGFR when age is ignored.

The correlation between marginal and age-specific genetic effects was high for 60-year-old (Spearman correlation coefficient, r=0.88; in line with mean age=57 years). The correlation was reduced for 50-year-old (r=0.71) and near zero for 70-year-old (r=0.06); the correlation between marginal and interaction effects was also near zero (r=0.04; **Figure 3c, d**). This causes the age-agnostic PGS to differ from the age-informative PGS for young and old individuals and from the interaction effect PGS.

### Age-informative PGS across 282 SNPs improves predictive ability in young and old individuals from UK Biobank and KORA&AugUR

In data from UK Biobank and KORA&AugUR, we generated the age-agnostic and age-informative PGSs and the main and interaction effect PGSs for the 282 kidney function SNPs (**Table 1**).

First, we were interested in how much of the eGFR variance was explained by the 282 SNPs and how much more the age-informative PGS explained compared to the age-agnostic PGS in cross-sectional analyses. Since we hypothesized that the age-informative PGS improved predictive ability in young and old individuals, we analyzed the datasets overall and by age-group (**Supplemental Figure S1, Supplemental Table S3**). Age groups were derived by integrating longitudinal data resulting in an age range of 25-78 age (UK Biobank n=254,068) or 25-98 years (KORA&AugUR n=9,001, age range: 25-98 years; **Table 1**). Over all individuals, the age-informative PGS explained slightly more of the eGFR variance than the age-agnostic PGS (UK Biobank: partial R^2^=5.5% versus 5.4%, KORA&AugUR: 3.9% versus 3.7%); in young and old individuals, the gain by the age-informative PGS was more pronounced (e.g. KORA&AugUR: young: 6.3% versus 5.9%, very-old: 2.3% versus 1.8%; **Figure 4a, Supplemental Tables S4&S5**). The two PGSs performed similar in middle-aged individuals, as expected. Total R^2^ underscored the strong predictive ability of age alone (UK Biobank: 20%; KORA&AugUR: 47%) with small improvement by adding the age-informative or the age-informative PGS (**Figure 4b**). In summary, this supported our idea that the age-informative PGS improved predictive ability compared to the age-agnostic PGS in young and old individuals. While the overall gain was small, the age-informative PGS always performed better.

**Figure 4.**
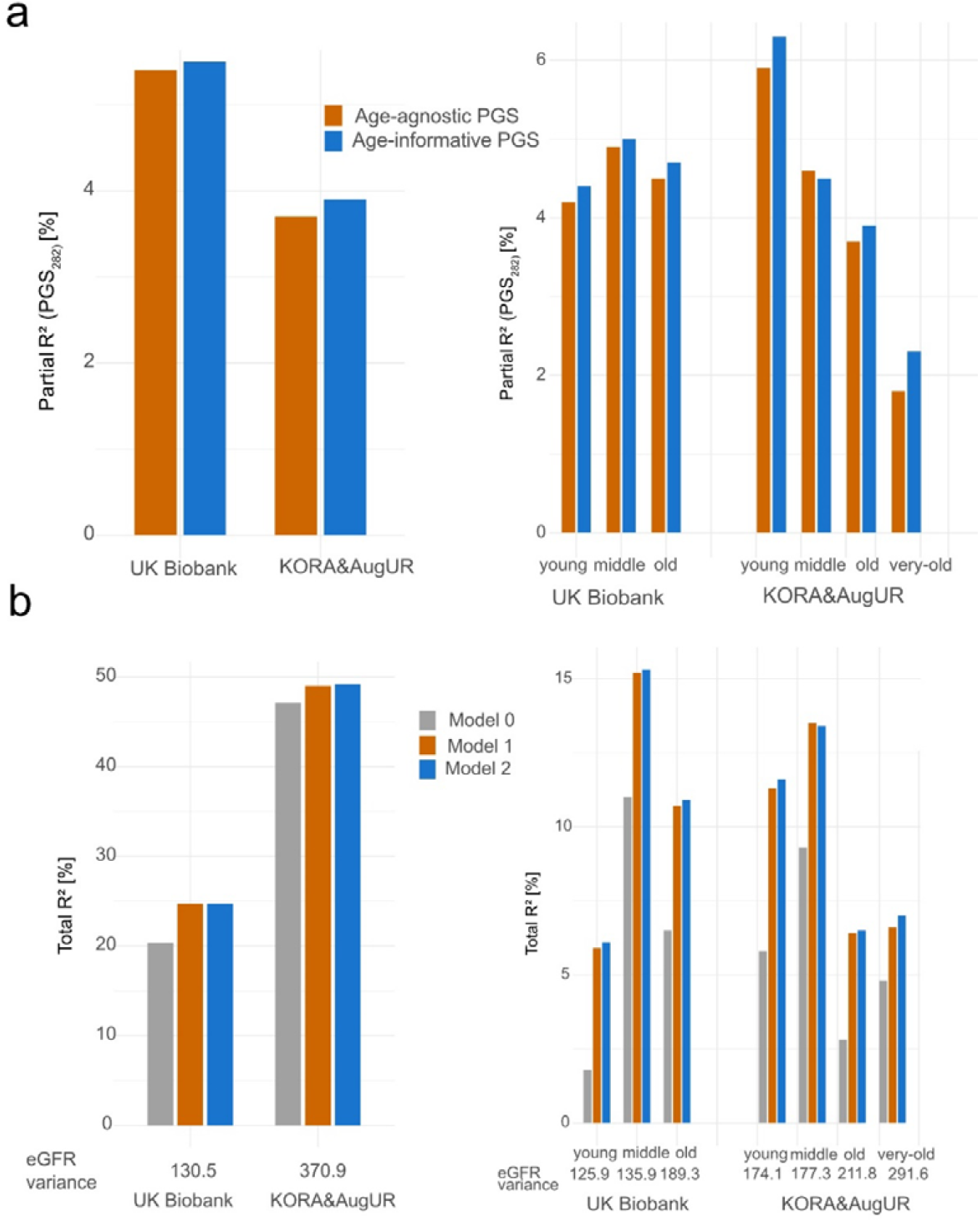
Improved predictive ability of the age-informative PGS versus age-agnostic PGS in young and old individuals. We fitted linear regression models on eGFR including i) age only (*model 0*), ii) age and age-agnostic PGS (*model 1*), or iii) age and age-informative PGS (*model 2*) in cross-sectional UK Biobank and KORA&AugUR. We evaluated the predictive ability by proportion of explained eGFR variance overall and in the different age groups (n= 41,099, n=276,096, n=115,103, n=254,084 for young, middle-aged, old aged, and overall in UK Biobank; n= 2,079, n=4,012, n=3,283, n= 2,807 for young, middle-aged, old aged, very-old-aged, and overall in KORA&AugUR). Shown are partial R^2^ of age-agnostic and age-informative PGS in the overall sample and by age groups and total R^2^ of the three models. The color code differentiates the PGSs or the three models. In (b), the eGFR variance is stated for each group in [ml/min/1.73m^2^]^2^. Detailed model results are provided in **Supplemental Tables S3 and S4**.

Second, we were interested in the predictive ability of our age-informative PGS (*model 2*) compared to the joint predictive ability of the main- and interaction-effect PGS in the model recommended by *Jayasinghe et al*. (*model 3*, again using cross-sectional data). The joint effect of the two separate PGSs showed the same predictive ability as our age-informative PGS (UK Biobank: partial R^2^=5.5%; KORA&AugUR: 3.9%; **Supplemental Table S6**). There was no difference between *model 3* or *model 4* by *Jagasinghe et al*. or between *model 1* and *model 1 extended* with a PGS*age interaction term (**Supplemental Table S6**). Thus, our age-informative PGS effectively combines the predictive ability of the two separate PGSs.

Third, when applying the corresponding LMMs to longitudinal data from UK Biobank (n=351,175, m=1,535,697) and KORA&AugUR (n=9,057, m=16,804), we found the pattern of the partial R^2^ across models to be similar compared to the results in cross-sectional data (**Supplemental Table S7** compared to **Supplemental Table S4&S6**). The slightly smaller R^2^ longitudinally versus cross-sectionally, particularly in UK Biobank, might be explained by more heterogeneous assessments of longitudinal eGFR-values (KORA&AugUR: different assays; UK Biobank: mainly from eHR, assays unknown). In summary, this underscored the improved performance by accounting for age in the PGS for eGFR and the applicability of the PGS in longitudinal as well as cross-sectional data.

### Interaction effect PGS improves predictive ability of eGFR-decline compared to conventional PGS

We postulated that the interaction effect PGS is a straight-forward approach for a PGS for eGFR-decline. We compared the interaction effect PGS with the age-agnostic PGS regarding association and predictive ability for eGFR-decline.

First, we compared association using results of *model 3L* versus *extended model 1L* in longitudinal data (UK Biobank, n=351,175, m=1,535,697; KORA&AugUR data, n=9,057, m=16,804; **Supplemental Table S7**): (i) the age effect provides mean annual decline in the reference group (50-year-old individuals with zero “bad” alleles): e.g. −0.28 or −1.02 mL/min/1.73m^2^ per year in KORA&AugUR in *model 3L* or *1L*, respectively. (ii) Association of the age-agnostic PGS with annual eGFR-decline was not significant (KORA&AugUR, 0.00078 mL/min/1.73m^2^ and allele, 95%-CI= −0.00080 to 0.00023, P=0.33). (iii) Association of the interaction effect PGS with annual eGFR-decline was statistically different from zero (P=1.5×10^−10^) and negative, i.e. each “bad” allele enhanced the annual decline (KORA&AugUR: −0.0018 mL/min/1.73m^2^ per allele, 95%-CI=−0.00025 to −0.00013). (iv) Results were similar between *models 3L* and *4L* and between KORA&AugUR and UK Biobank, except the age-agnostic PGS was significantly different from zero in UK Biobank.

Second, we analyzed the PGS association with eGFR-decline in the two-step approach (person-specific slopes). The interaction effect PGS increased the R^2^ compared to the age-agnostic PGS: in UK Biobank to R^2^=0.3% versus 0.2% and in KORA&AugUR to 1.0% versus 0.7% (**Supplemental Table S8**). We observed a smaller effect (in absolute value) of the interaction effect PGS in the two-step approach compared to *model 3L* (**Supplemental Table S8**, e.g. KORA&AugUR, −0.00075 versus −0.0018 mL/min/1.73m^2^ per allele and year). This is in line with the known bias in risk factor association using the two-step approach ^16, 31^. In summary, these results underscored that the interaction effect PGS performed better than the age-agnostic PGS to predict eGFR-decline. While the PGS predictive ability for eGFR-decline was small, it should be noted that this reflected the prediction “from the cradle” (only genetics) irrespective of acquired (“non-genetic”) risk factors like diabetes, obesity or albuminuria.

### Genetic profiles to predict eGFR and eGFR-decline on population- and individual-level

Finally, we evaluated whether genetic profiles could differentiate future eGFR values for persons at a certain age or future eGFR-decline on the population- and individual-level. We used KORA&AugUR cross-sectional (for eGFR, n=9,001) and longitudinal (for eGFR-decline, n=9,057, m=16,804).

First, we examined extreme genetic profiles of the age-informative PGS (using *model 2* results to predict eGFR, **Supplemental Table S4**) or the interaction effect PGS (using *model 3L* results to predict eGFR-decline; **Supplemental Table S7**). When comparing individuals with 200 versus 364 “bad” alleles, average eGFR was 120 versus 78.7 mL/min/1.73m^2^ for a 40-year-old and 90.5 versus 49.1 mL/min/1.73m^2^ for an 80-year-old individual; average eGFR-decline was −0.65 versus −0.95 mL/min/1.73m^2^ per year (**Table 2**). Corresponding 95%-CIs were small and distinct between profiles, indicating high estimation precision and effective differentiation of mean eGFR and eGFR-decline by genetic profiles on the population-level. On the individual level, we judged the 95%-prediction intervals: these were very wide and overlapping across profiles.

**Table 2.**
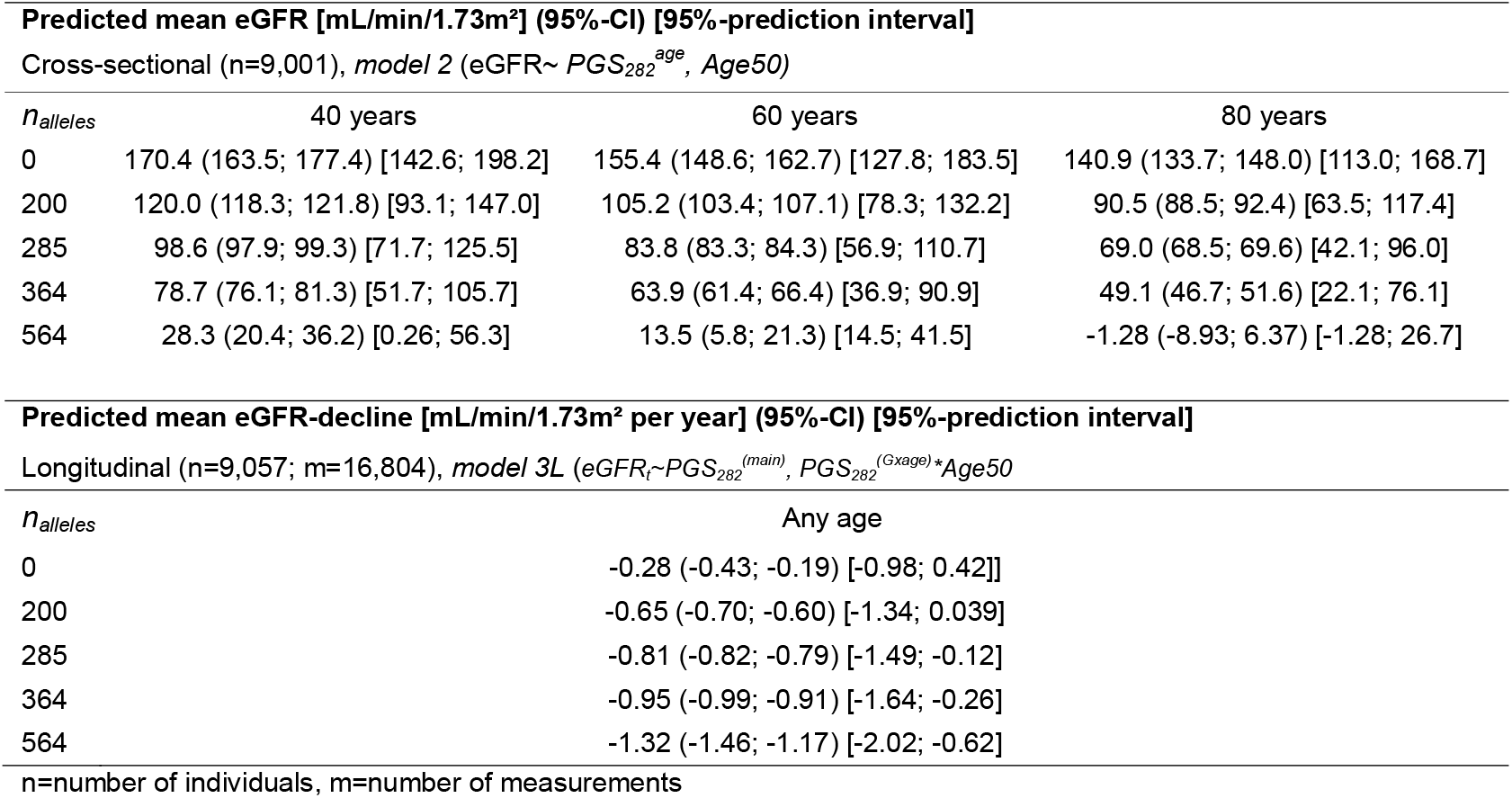
Prediction of eGFR-levels and eGFR-decline across different genetic profiles in KORA&AugUR. We evaluated the age-informative PGS, (PGS_282_^age^) based on 282 kidney function SNPs for individuals with 40, 50, 60, 70, 80 years of age in KORA&AugUR. We used results from a linear regression on eGFR including age and PGS in cross-sectional data (*model 2*, **Supplemental Table S4**). We predicted eGFR values by age and genetic profiles with various numbers of eGFR-lowering alleles 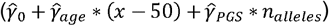. Shown are predicted mean eGFR with 95%-CI and 95%-prediction intervals. We also derived the interaction effect PGS (PGS_282_^Gxage^) in KORA&AugUR. Based on *model 3L* applied in longitudinal data (**Supplemental Table S7**), we predicted eGFR-decline for various numbers of eGFR-lowering alleles (*n*_*alleles*_) combining the age effect with the interaction effect 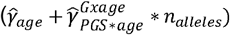. Shown is the estimated mean annual eGFR-decline with respective 95%-CIs and 95%-prediction intervals.

| Predicted mean eGFR [mL/min/1.73m²] (95%-CI) [95%-prediction interval] |  |  |  |  |
| --- | --- | --- | --- | --- |
| Cross-sectional (n=9,001), *model 2* (eGFR~ $PGS_{282}^{age}$, Age50) | | | | |
| $n_{alleles}$ | 40 years | 60 years | 80 years | |
| 0 | 170.4 (163.5; 177.4) [142.6; 198.2] | 155.4 (148.6; 162.7) [127.8; 183.5] | 140.9 (133.7; 148.0) [113.0; 168.7] |  |
| 200 | 120.0 (118.3; 121.8) [93.1; 147.0] | 105.2 (103.4; 107.1) [78.3; 132.2] | 90.5 (88.5; 92.4) [63.5; 117.4] |  |
| 285 | 98.6 (97.9; 99.3) [71.7; 125.5] | 83.8 (83.3; 84.3) [56.9; 110.7] | 69.0 (68.5; 69.6) [42.1; 96.0] |  |
| 364 | 78.7 (76.1; 81.3) [51.7; 105.7] | 63.9 (61.4; 66.4) [36.9; 90.9] | 49.1 (46.7; 51.6) [22.1; 76.1] |  |
| 564 | 28.3 (20.4; 36.2) [0.26; 56.3] | 13.5 (5.8; 21.3) [14.5; 41.5] | -1.28 (-8.93; 6.37) [-1.28; 26.7] |  |
| Predicted mean eGFR-decline [mL/min/1.73m² per year] (95%-CI) [95%-prediction interval] |  |  |  |  |
| Longitudinal (n=9,057; m=16,804), *model 3L* (eGFR<sub>t</sub>~$PGS_{282}^{(main)}$, $PGS_{282}^{(G \times age)} \times Age50$) | | | | |
| $n_{alleles}$ | Any age | | | |
| 0 | -0.28 (-0.43; -0.19) [-0.98; 0.42]] |  |  |  |
| 200 | -0.65 (-0.70; -0.60) [-1.34; 0.039] |  |  |  |
| 285 | -0.81 (-0.82; -0.79) [-1.49; -0.12] |  |  |  |
| 364 | -0.95 (-0.99; -0.91) [-1.64; -0.26] |  |  |  |
| 564 | -1.32 (-1.46; -1.17) [-2.02; -0.62] |  |  |  |
| n=number of individuals, m=number of measurements |  |  |  |  |

Second, when comparing high-versus low-risk groups (highest versus lowest quintile), average eGFR differed by about 8 mL/min/1.73m^2^ at all ages (*model 2Q*, **Figure 5a**); average eGFR-decline was −0.75 mL/min/1.73m^2^ versus −0.88 per year (*model 3Q*; **Figure 5b, Supplemental Table S9**). Again, 95%-CIs indicated effective differentiation, while 95%-prediction intervals were wide and undistinguishable between genetic risk groups. Thus, the stratification by high versus low PGS quintiles can distinguish eGFR and eGFR-decline on the population level, but not on the individual level.

**Figure 5.**
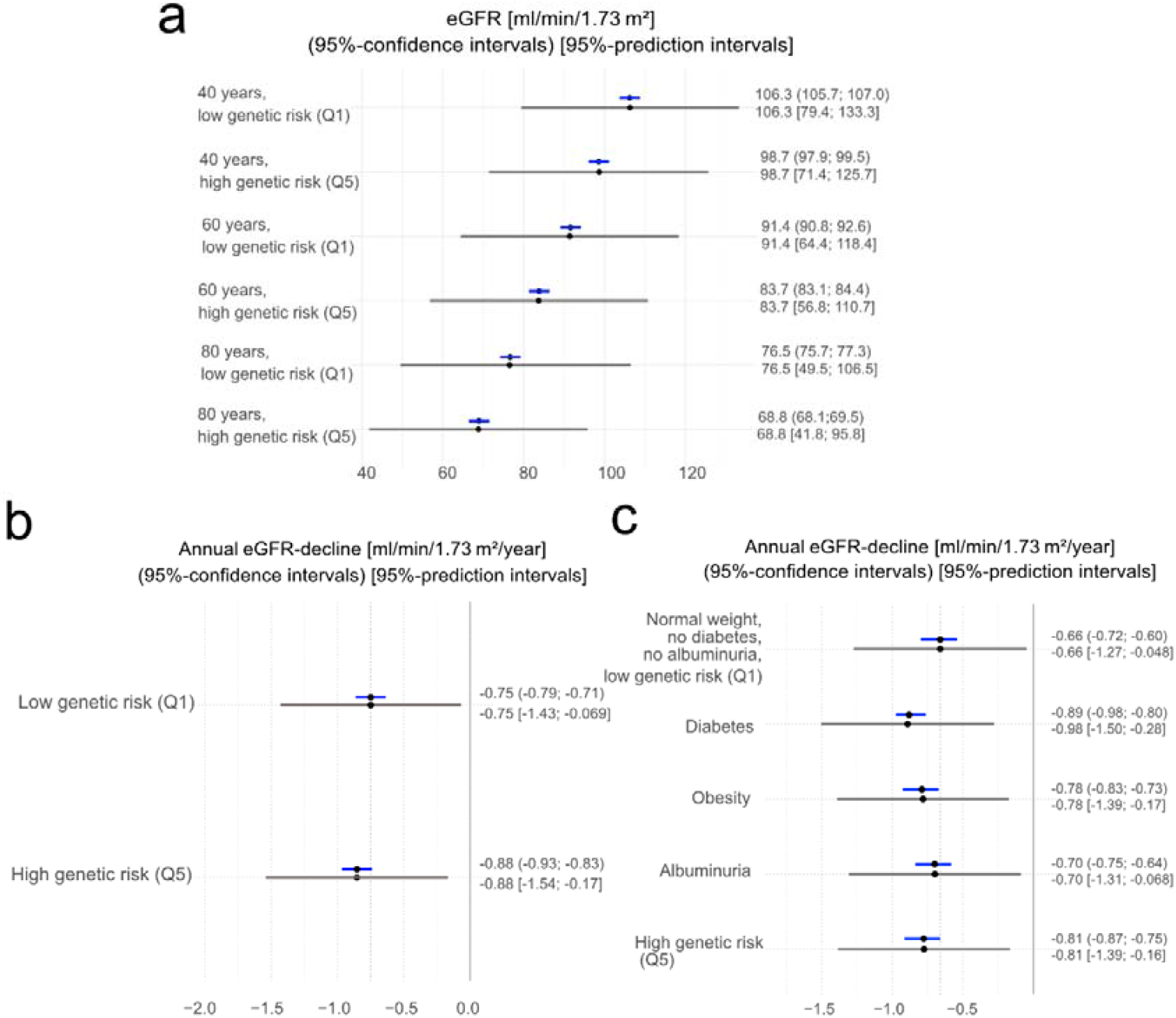
Comparison of genetic and non-genetic risk groups to predict eGFR and eGFR-decline on population- and individual-level. We derived high and low genetic risk groups using the highest and lowest quintile of the age-informative PGS and the interaction effect PGS (Q1, Q5). We applied linear regression with the age-informative PGS quintiles cross-sectional data (*model 2Q*) and LMM with the interaction effect PGS and its interaction with age in longitudinal data (*model 3Q*) in KORA&AugUR (n=9,001 or n=9,057 and m=16,804, respectively). Shown are (**a**) mean values of eGFR by age and genetic risk group (Q1, Q5) and (**b**) mean values of eGFR-decline by genetic risk group (interaction effect PGS Q1 and Q5) and corresponding 95%-CIs (blue) and 95%-prediction intervals (grey). The *model 3Q* was then extended to include non-genetic risk factors (diabetes, obesity, albuminuria) and their interactions with age and applied in KORA&AugUR (n=7,988; m=10,742; **Supplemental Table S9**). Shown is (**c**) the mean eGFR-decline by genetic and nongenetic risk groups and corresponding 95%-CIs (blue) and 95%-prediction intervals (grey).

Finally, we put the high versus low genetic risk stratification for eGFR-decline into perspective to the risk stratification by acquired risk factors (diabetes, obesity, and albuminuria). Adding these risk factors and their interaction with age into *model 3Q* showed similar average eGFR-decline of the high genetic risk group as for diabetes, obesity or albuminuria (**Figure 5c, Supplemental Table S9**). The 95%-prediction intervals were wide and overlapping across all groups. This underscores that genetic risk strata differentiate eGFR-decline on the population-level in a similar fashion as the status of diabetes, obesity, or albuminuria, but also that a person’s eGFR-decline remains difficult to predict with any of these factors.

## Discussion

We introduced a concept for an age-informative PGS that improved predictive ability for quantitative traits where genetic effects depend on age. We documented its applicability and meaningful differentiation of predicted kidney function and kidney function decline on the population-level in a study that covered an age range from 25 to 98 years of age.

Regarding methodology, we showed better predictive ability of our age-informative PGS than a conventional age-agnostic PGS in theory and in real data of a 282-SNP PGS for kidney function. The improved performance was particularly pronounced for young and old individuals. This implicates that predictive ability of a PGS is underestimated in young and old individuals when age is ignored. We also showed that our age-informative PGS effectively combined the predictive ability of two PGSs generated with the main effects or interaction effects separately, which was suggested previously to account for GxE^5^. Our age-informative PGS thus provides a simple, general approach for a PGS that encapsulates GxE effects into a single, interpretable score. For clinical application, a single score is more practical to use for risk stratification and easier to interpret. Still, the separation of the PGS into main effect and interaction effect PGSs analyzed in the model recommended by *Jayasinghe et al*. enabled an interesting novel application: the interaction effect PGS is a straight-forward to approach for a PGS for trait change over age. We showed that the interaction effect PGS improved predictive ability for kidney function decline compared to the conventional PGS based on marginal genetic effects. Since trait change is often highly relevant clinically, as this is the case for kidney function decline, this fills a gap in methodology that is clinically important.

Important for personalized medicine are our findings of the genetic prediction of kidney function and its decline over age. Our results showed that stratification by PGS quintiles can distinguish eGFR and eGFR-decline meaningfully on the population level and to a similar extent as stratification by acquired risk factors like diabetes, obesity or albuminuria. This implicates that measures to “remove” the risk factor would have the same impact on the population. This is with the caveat that such measures are difficult to envision regarding an unfavorable genetic profile, while the “removal” of high glucose is available by anti-diabetic medication. The prediction of kidney function decline on the individual level is challenging due to the high variability of random slopes^20^. Our results underscore that neither genetic risk nor acquired risks like diabetes, obesity or albuminuria help differentiate eGFR-decline on the individual level. Thus, it remains difficult to predict eGFR-decline with any of these risk factors, genetic or acquired.

There has been little previous work regarding the predictive ability of a PGS for eGFR. Our previous score of 594 SNPs known for creatinine-based eGFR association had shown an R^2^ of 9.3% in UK Biobank^15^ and 4.6% in AugUR^32^. The smaller proportion of the trait variance explained by the 594-variant PGS in AugUR compared to UK Biobank was due to a larger eGFR variance in old individuals. That work included SNPs with effects on creatinine metabolism rather than kidney function, which were removed here by restriction to 282 SNPs ascertained with cystatin-based eGFR association. While age-agnostic PGS for eGFR have been evaluated in the context of kidney failure and CKD progression (e.g.^33,34^), to our knowledge, no prior study has examined the effect of a PGS on eGFR decline. There are prediction models for eGFR-decline based on “non-genetic” factors that documented HbA1c, BMI, and UACR as the most pronounced risk factors for accelerated decline, but each with small effects^35,12,36,37^, in line with our findings. Comparing those results with ours more quantitatively is challenging, since eGFR is modelled over time and 95% prediction intervals are not provided. We model eGFR over age and recommend 95% prediction intervals as an intuitive range of values to help judge the individual-level eGFR-decline, which is the eGFR-decline to be expected in 95% of the underlying population.

The concept of genetic effects on “trait change over age”, like eGFR-decline, is equivalent to “age-dependent genetic effects” for the trait under certain assumptions^38^. A large body of literature is about age-dependency of genetic effects for diseases like Alzheimer or prostate cancer, where age diminishes genetic effects probably due to increasingly accumulating acquired risks^8, 7^. Genetic effects that become more pronounced by age appear to be rather specific to kidney function^16^ and blood pressure traits^38^. Kidney function is thus an ideal role model to evaluate the impact when accounting for age in the PGS. Our PGS can account for age-dependent genetic effects when the underlying GWAS models interactions with age as done in *Wiegrebe et al*., while simple age adjustment in GWAS removes a mediating effect of age in the genetic association. We demonstrated that genetic effects on kidney function were overestimated in young and underestimated for old individuals when age-dependency was ignored. For average-aged individuals, agnostic (marginal) effects work well, since they average over all individuals in the dataset.

Some strengths and limitations should be noted. While we were able to show improved prediction by the age-informative PGS compared to the age-agnostic PGS, the overall gain in R^2^ in real data was small. Still, the application is simple and there is thus no good reason to keep using the age-agnostic PGS when genetic effects for a trait depend on age. The limited age range in the longGWAS used to derive age-specific genetic effects for eGFR, which was 25 to 78 years in the longitudinal UK Biobank dataset, required us to extrapolate age-specific effects for individuals with > 80 years of age. Old-age individuals are typically underrepresented in the GWAS of population-based studies, since elderly require a study program tailored to their needs. We had the opportunity to integrate AugUR with KORA, as established before^20^, which provided a validation dataset with good representation of elderly. Future larger GWAS accounting for age and integrating a wide age range in the search for SNP-effects will improve the data basis. One limitation is the assumption of a linear relationship of age on the SNP-effect that might have left non-linear patterns unidentified. It is a strength of our approach of age-specific weights, since such weights can also be derived from non-linear models. Future work might also evaluate extended approaches for the selection of SNPs into the PGS by using different significance thresholds (e.g. LDpred^39^ or PRSice^40^), but this requires larger longGWAS and larger validation data. While we excluded SNPs associated solely with creatinine metabolism rather than kidney function, future work could further integrate cystatin C measurements to derive eGFR more comprehensively.^27^ in GWAS and validation data. Our analyses were limited to individuals of European ancestry, and the findings may therefore not be directly generalizable to populations of other ancestries. The present work focuses on optimizing the PGS itself, a necessary step before assessing its potential clinical utility. Further studies are needed to evaluate the ability of age-informative PGSs to help predict individuals at high risk for CKD onset and progression.

An important strength of our approach to address GxE in PGS is the generalizability to other quantitative traits and SNP-interactions with other quantitative factors, such as physical activity or frequency of alcohol consumption^41^. Another strength is the analysis of two datasets, UK Biobank as the weight-identifying data and KORA&AugUR as a completely independent dataset, which demonstrated comparable performance despite some expectation that performance would be higher in the weight-identifying data. One reason can be the use of SNPs with significant associations rather than a genome-wide approach to generate the PGS. Given that GWAS are typically age-adjusted without SNP interaction by age, the example of kidney function genetics emphasizes the utility of age interaction terms. Our concept of an age-informative PGS is complementary to the model that has been proposed by *Jayasinghe et al*. to account for GxE interactions using the main effect PGS and interaction effect PGS^5^. We thus recommend the use of both approaches. For clinical application, a single score is more practical for risk stratification for eGFR. For eGFR-decline risk stratification with the interaction effect PGS, the *Jayasinghe* model is very intuitive. We showed that our both approaches are applicable in cross-sectional as well as longitudinal data.

In conclusion, our age-informative PGS approach is applicable to quantitative traits with general GxE and fills an important gap for an approach that is simple to apply. Kidney function served here as a role model due to its strong age-dependency of genetic effects. While the prediction of kidney function and kidney function decline by genetic risk strata remains difficult on the individual level, the relevance of the genetic profile on the population-level prompts further work to understand the underlying mechanisms.

## Acknowledgements

This work was funded by the Deutsche Forschungsgemeinschaft (DFG, German Research Foundation) – Project-ID 387509280, SFB1350 (Subproject C6 to I.M.H.) and Project-ID 509149993, TRR 374. We conducted this research using the UK Biobank resource under the application number 20272. The AugUR study was supported by the German Federal Ministry of Education and Research (grant number BMBF 01ER1206, BMBF 01ER1507 to I.M.H.) and by the German Research Foundation (grant number HE 3690/7-1 to I.M.H). The KORA study was initiated and financed by the Helmholtz Zentrum München - German Research Center for Environmental Health, which is funded by the German Federal Ministry of Education and Research (BMBF) and by the State of Bavaria. Data collection in the KORA study is done in cooperation with the University Hospital of Augsburg.

We would like to thank all study participants for taking part in the studies. We further thank the physicians and health insurance companies supporting the studies and all study nurses for their expert work in performing the study visits.

## Author Contributions

JMH performed statistical analyses and manuscript writing; SW: supported statistical analyses and interpretation of results; TWW: Statistical expertise, GWAS on eGFR_cys_; MG: UK Biobank data preparation; B.T. and C.G. provided data for the KORA study; FH: statistical expertise; AP: study PI (KORA); HK: statistical expertise and interpretation of results; IMH: was the study PI, initiated the project, supervised statistical analyses, and designed and wrote the manuscript. All authors contributed to the reviewing and editing of the manuscript and approved the final version.

## Ethics approval

This UKB project was conducted under the application number 20272. The AugUR study was approved by the Ethics Committee of the University of Regensburg, Germany (vote 12-101-0258). The KORA-S3 study was approved by the local authorities and conducted in accordance with the data protection regulations as part of the World Health Organization Monitoring Trends and Determinants in Cardiovascular Disease (MONICA) Project. All other KORA studies were approved by the Ethics Committee of the Bavarian Chamber of Physicians (KORA-F3 EC Number 03097, KORA-S4 EC Number 99186, KORA-F4/FF4 EC Number 06068, KORA-Fit EC Number 17040). All studies comply with the 1964 Declaration of Helsinki and its later amendments. All participants provided written informed consent.

## Data and code availability

This UK Biobank project was conducted under the application number 20272. UK Biobank is a publicly accessible database. Individual participant data from UKB are available via the UK Biobank resource. Individual participant data from KORA-3, KORA-4, and AugUR are not publicly available due to data protection regulations and restrictions imposed by the Ethics Committee of the Bavarian Chamber of Physicians to protect participant privacy. However, data can be accessed upon request through project agreements with KORA (https://helmholtz-muenchen.managed-otrs.com/external) or AugUR. We provide summary statistics for kidney function genetic variant associations (see Supplemental Data 1). Original data for the eGFR associated genetic variants are publicly available (Wiegrebe et al., 2024; DOI:10.1038/s41467-024-54483-9). The code to generate age-informative PGS, is prepared for publication and will be available on our GitHub repository (<u>Genetic Epidemiology Regensburg · GitHub</u>).

## References

1. Khera, A.V., Chaffin, M., Aragam, K.G., Haas, M.E., Roselli, C., Choi, S.H., Natarajan, P., Lander, E.S., Lubitz, S.A., and Ellinor, P.T., et al. (2018). Genome-wide polygenic scores for common diseases identify individuals with risk equivalent to monogenic mutations. Nature genetics 50, 1219–1224.

2. Torkamani, A., Wineinger, N.E., and Topol, E.J. (2018). The personal and clinical utility of polygenic risk scores. Nature reviews. Genetics 19, 581–590.

3. Dudbridge, F. (2013). Power and predictive accuracy of polygenic risk scores. PLoS genetics 9, e1003348.

4. Rao, D.C., Sung, Y.J., Winkler, T.W., Schwander, K., Borecki, I., Cupples, L.A., Gauderman, W.J., Rice, K., Munroe, P.B., and Psaty, B.M. (2017). Multiancestry Study of Gene-Lifestyle Interactions for Cardiovascular Traits in 610 475 Individuals From 124 Cohorts: Design and Rationale. Circulation. Cardiovascular genetics 10.

5. Jayasinghe, D., Momin, M.M., Beckmann, K., Hyppönen, E., Benyamin, B., and Lee, S.H. (2024). Mitigating type 1 error inflation and power loss in GxE PRS: Genotype-environment interaction in polygenic risk score models. Genetic epidemiology 48, 85– 100.

6. Yang Xu, C., Gupte, T., Hoffmann, T.J., Iribarren, C., Zhou, X., and Ganesh, S.K. (2024). Sex-specific genetic architecture of blood pressure. Nature medicine 30, 818– 828.

7. Schaid, D.J., Sinnwell, J.P., Batzler, A., and McDonnell, S.K. (2022). Polygenic risk for prostate cancer: Decreasing relative risk with age but little impact on absolute risk. American journal of human genetics 109, 900–908.

8. Escott-Price, V., and Schmidt, K.M. (2023). Pitfalls of predicting age-related traits by polygenic risk scores. Annals of human genetics 87, 203–209.

9. (2024). KDIGO 2024 Clinical Practice Guideline for the Evaluation and Management of Chronic Kidney Disease. Kidney international 105, S117–S314.

10. Denic, A., Lieske, J.C., Chakkera, H.A., Poggio, E.D., Alexander, M.P., Singh, P., Kremers, W.K., Lerman, L.O., and Rule, A.D. (2017). The Substantial Loss of Nephrons in Healthy Human Kidneys with Aging. Journal of the American Society of Nephrology: JASN 28, 313–320.

11. Wetzels, J.F.M., Willems, H.L., and Heijer, M.den (2008). Age- and gender-specific reference values of estimated glomerular filtration rate in a Caucasian population: Results of the Nijmegen Biomedical Study. Kidney international 73, 657–658.

12. Gregorich, M., Kammer, M., Heinzel, A., Böger, C., Eckardt, K.U., Heerspink, H.L., Jung, B., Mayer, G., Meiselbach, H., and Schmid, M., et al. (2023). Development and Validation of a Prediction Model for Future Estimated Glomerular Filtration Rate in People With Type 2 Diabetes and Chronic Kidney Disease. JAMA network open 6, e231870.

13. Baba, M., Shimbo, T., Horio, M., Ando, M., Yasuda, Y., Komatsu, Y., Masuda, K., Matsuo, S., and Maruyama, S. (2015). Longitudinal Study of the Decline in Renal Function in Healthy Subjects. PloS one 10, e0129036.

14. Wuttke, M., Li, Y., Li, M., Sieber, K.B., Feitosa, M.F., Gorski, M., Tin, A., Wang, L., Chu, A.Y., and Hoppmann, A., et al. (2019). A catalog of genetic loci associated with kidney function from analyses of a million individuals. Nature genetics 51, 957–972.

15. Stanzick, K.J., Li, Y., Schlosser, P., Gorski, M., Wuttke, M., Thomas, L.F., Rasheed, H., Rowan, B.X., Graham, S.E., and Vanderweff, B.R., et al. (2021). Discovery and prioritization of variants and genes for kidney function in 1.2 million individuals. Nature Communications 12, 4350.

16. Wiegrebe, S., Gorski, M., Herold, J.M., Stark, K.J., Thorand, B., Gieger, C., Böger, C.A., Schödel, J., Hartig, F., and Chen, H., et al. (2024). Analyzing longitudinal trait trajectories using GWAS identifies genetic variants for kidney function decline. Nature Communications 15, 10061.

17. Gorski, M., Wiegrebe, S., Burkhardt, R., Behr, M., Küchenhoff, H., Stark, K.J., Böger, C.A., and Heid, I.M. (2025). Bias-corrected serum creatinine from UK Biobank electronic medical records generates an important data resource for kidney function trajectories. Scientific reports 15, 3540.

18. Holle, R., Happich, M., Löwel, H., and Wichmann, H.E. (2005). KORA--a research platform for population based health research. Gesundheitswesen (Bundesverband der Arzte des Offentlichen Gesundheitsdienstes (Germany)) 67 Suppl 1, S19–25.

19. Stark, K., Olden, M., Brandl, C., Dietl, A., Zimmermann, M.E., Schelter, S.C., Loss, J., Leitzmann, M.F., Böger, C.A., and Luchner, A., et al. (2015). The German AugUR study: study protocol of a prospective study to investigate chronic diseases in the elderly. BMC geriatrics 15, 130.

20. Herold, J.M., Wiegrebe, S., Nano, J., Jung, B., Gorski, M., Thorand, B., Koenig, W., Zeller, T., Zimmermann, M.E., and Burkhardt, R., et al. (2024). Population-based reference values for kidney function and kidney function decline in 25-to 95-year-old Germans without and with diabetes. Kidney international 106, 699–711.

21. Inker, L.A., Eneanya, N.D., Coresh, J., Tighiouart, H., Wang, D., Sang, Y., Crews, D.C., Doria, A., Estrella, M.M., and Froissart, M., et al. (2021). New Creatinine- and Cystatin C-Based Equations to Estimate GFR without Race. The New England journal of medicine 385, 1737–1749.

22. Walter, K., Min, J.L., Huang, J., Crooks, L., Memari, Y., McCarthy, S., Perry, J.R.B., Xu, C., Futema, M., and Lawson, D., et al. (2015). The UK10K project identifies rare variants in health and disease. Nature 526, 82–90.

23. McCarthy, S., Das, S., Kretzschmar, W., Delaneau, O., Wood, A.R., Teumer, A., Kang, H.M., Fuchsberger, C., Danecek, P., and Sharp, K., et al. (2016). A reference panel of 64,976 haplotypes for genotype imputation. Nature genetics 48, 1279–1283.

24. Wichmann, H.-E., Gieger, C., and Illig, T. (2005). KORA-gen--resource for population genetics, controls and a broad spectrum of disease phenotypes. Gesundheitswesen (Bundesverband der Arzte des Offentlichen Gesundheitsdienstes (Germany)) 67 Suppl 1, S26–30.

25. Herold, J.M., Nano, J., Gorski, M., Winkler, T.W., Stanzick, K.J., Zimmermann, M.E., Brandl, C., Peters, A., Koenig, W., Burkhardt, R., Gessner, A., Heid, I.M., Gieger, C. and Stark, K.J. (2023). Polygenic scores for estimated glomerular filtration rate in a population of general adults and elderly – comparative results from the KORA and AugUR study, https://bmcgenomdata.biomedcentral.com/articles/10.1186/s12863-023-01130-9.

26. Stanzick, K.J., Stark, K.J., Gorski, M., Schödel, J., Krüger, R., Kronenberg, F., Warth, R., Heid, I.M., and Winkler, T.W. (2023). KidneyGPS: a user-friendly web application to help prioritize kidney function genes and variants based on evidence from genome-wide association studies. BMC bioinformatics 24, 355.

27. Inker, L.A., Schmid, C.H., Tighiouart, H., Eckfeldt, J.H., Feldman, H.I., Greene, T., Kusek, J.W., Manzi, J., van Lente, F., and Zhang, Y.L., et al. (2012). Estimating glomerular filtration rate from serum creatinine and cystatin C. The New England journal of medicine 367, 20–29.

28. Fahrmeir, L., Kneib, T., and Lang, S. (2009). Regression (Berlin, Heidelberg: Springer Berlin Heidelberg).

29. Koraishy, F.M., Mann, F.D., Waszczuk, M.A., Kuan, P.-F., Jonas, K., Yang, X., Docherty, A., Shabalin, A., Clouston, S., and Kotov, R., et al. (2022). Polygenic association of glomerular filtration rate decline in world trade center responders. BMC nephrology 23, 347.

30. WHO Collaborating Centre for Drug Statistics Methodology (2012). Guidelines for ATC classification and DDD assignment 2013.

31. Hastie, T., Tibshirani, R., and Friedman, J.H. (2009). The elements of statistical learning. Data mining, inference, and prediction (New York, NY: Springer).

32. Herold, J.M., Nano, J., Gorski, M., Winkler, T.W., Stanzick, K.J., Zimmermann, M.E., Brandl, C., Peters, A., Koenig, W., and Burkhardt, R., et al. (2023). Polygenic scores for estimated glomerular filtration rate in a population of general adults and elderly - comparative results from the KORA and AugUR study. BMC genomic data 24, 28.

33. Steinbrenner, I., Yu, Z., Jin, J., Schultheiss, U.T., Kotsis, F., Grams, M.E., Coresh, J., Wuttke, M., Kronenberg, F., and Eckardt, K.-U., et al. (2023). A polygenic score for reduced kidney function and adverse outcomes in a cohort with chronic kidney disease. Kidney international 103, 421–424.

34. Khan, A., Turchin, M.C., Patki, A., Srinivasasainagendra, V., Shang, N., Nadukuru, R., Jones, A.C., Malolepsza, E., Dikilitas, O., and Kullo, I.J., et al. (2022). Genome-wide polygenic score to predict chronic kidney disease across ancestries. Nature medicine 28, 1412–1420.

35. Schaeffner, E.S., Ebert, N., Kuhlmann, M.K., Martus, P., Mielke, N., Schneider, A., van der Giet, M., and Huscher, D. (2022). Age and the Course of GFR in Persons Aged 70 and Above. Clinical journal of the American Society of Nephrology: CJASN 17, 1119– 1128.

36. Waas, T., Schulz, A., Lotz, J., Rossmann, H., Pfeiffer, N., Beutel, M.E., Schmidtmann, I., Münzel, T., Wild, P.S., and Lackner, K.J. (2021). Distribution of estimated glomerular filtration rate and determinants of its age dependent loss in a German population-based study. Scientific reports 11, 10165.

37. Grams, M.E., Brunskill, N.J., Ballew, S.H., Sang, Y., Coresh, J., Matsushita, K., Surapaneni, A., Bell, S., Carrero, J.J., and Chodick, G., et al. (2022). Development and Validation of Prediction Models of Adverse Kidney Outcomes in the Population With and Without Diabetes. Diabetes care 45, 2055–2063.

38. Winkler, T.W., Wiegrebe, S., Herold, J.M., Stark, K.J., Küchenhoff, H., and Heid, I.M. (2024). Genetic-by-age interaction analyses on complex traits in UK Biobank and their potential to identify effects on longitudinal trait change. Genome biology 25, 300.

39. Privé, F., Arbel, J., and Vilhjálmsson, B.J. (2021). LDpred2: better, faster, stronger. Bioinformatics (Oxford, England) 36, 5424–5431.

40. Choi, S.W., and O’Reilly, P.F. (2019). PRSice-2: Polygenic Risk Score software for biobank-scale data. GigaScience 8.

41. Rask-Andersen, M., Karlsson, T., Ek, W.E., and Johansson, Å. (2017). Gene-environment interaction study for BMI reveals interactions between genetic factors and physical activity, alcohol consumption and socioeconomic status. PLoS genetics 13, e1006977.

