## Supplementary Material for "A novel age-informative polygenic score improves predictive ability for kidney function and kidney function decline"

### for


| | |
|---|---|
| Supplemental Note 1 | Relationship between our age-informative PGS and the joint main and interaction effect PGS |
| Supplemental Figure S1 | Differences and similarities of the distribution for age-agnostic and age-informative PGS across 282 kidney function SNPs in UK Biobank and KORA&AugUR |
| Supplemental Table S1 | Weights for unscaled and scaled PGS in the theoretical example |
| Supplemental Table S2 | Age-specific beta-estimates for 12 age-dependent SNPs in UK Biobank |
| Supplemental Table S3 | Descriptive statistics of the age-agnostic and age-informative PGS by study and age group |
| Supplemental Table S4 | Performance of age-agnostic and age-informative PGS for kidney function in cross-sectional data |
| Supplemental Table S5 | Performance of age-agnostic and age-informative PGS for kidney function by age group in cross-sectional data |
| Supplemental Table S6 | Performance of main and interaction-effects PGS for kidney function in *Jayasinghe* models |
| Supplemental Table S7 | Comparison of the predictive ability for kidney function of the PGSs in longitudinal data |
| Supplemental Table S8 | Predictive ability of age-agnostic and interaction-effect PGS for eGFR- decline using person-specific slopes in a two-step approach |
| Supplemental Table S9 | Association of genetic and non-genetic risk factors with change of eGFR over age |
| Supplemental References | |

**Supplemental Note 1. Relationship between our age-informative PGS and the joint main and interaction effect PGS**

We here describe the relationship between our age-informative PGS and the joint view of the main effect PGS and the interaction PGS. To reduce complexity, we first consider PGSs with unscaled weights. The age-informative PGS for an individual at x years of age is calculated as $PGS_k^x = \sum_{j=1}^{k} w_j^{(age=x)} * G_j$, where weights are age-specific genetic effect, $w_j^{(age=x)} := \beta_j^{(age=x)} := |\hat{\beta}_{G_j}^{(int)} + \hat{\beta}_{G_j*age}^{(int)} * (x-50)|$. The effect allele that is counted in the PGS is the eGFR-lowering allele based on the marginal effect under the idea that this is "bad" allele for eGFR in the population. When AGE is centered at 50 years in the analysis to derive the main effect, the main effect, $\hat{\beta}_{G_j}^{(int)}$, is the genetic effect on eGFR at 50 years of age. We assume that $\hat{\beta}_{G_j}^{(int)}$<0 for all SNPs and that the interaction effect, $\hat{\beta}_{G_j*age}^{(int)}$, is <0 for most SNPs (i.e. the eGFR-decline accelerating allele is the eGFR-lowering allele in the population). Then $\beta_j^{(age=x)}$<0 for most SNPs at all ages in the age spectrum of a dataset (assume age= 50 to 80 years) and the sum of these is negative. The sum in the age-informative PGS, $PGS_k^x = \sum_{j=1}^{k}(|\hat{\beta}_{G_j}^{(int)} + \hat{\beta}_{G_j*age}^{(int)} * (x-50)| * G_j)$ can then be separated into a sum of main effects and a sum of interaction effects,

$$\sum_{j=1}^{k} |\hat{\beta}_{G_j}^{(int)}| * G_j + (\sum_{j=1}^{k}(|\hat{\beta}_{G_j*age}^{(int)} * (x-50)| * G_j),$$

since both summands are negative. This can also be written as

$$= \sum_{j=1}^{k} |\hat{\beta}_{G_j}^{(int)}| * G_j + (\sum_{j=1}^{k}(|\hat{\beta}_{G_j*age}^{(int)}| * G_j * (x-50)$$

which corresponds to $PGS^{main} + PGS^{Gxage} * (x-50)$. This shows the relationship of our age-informative PGS with the main and interaction effect PGS of a person at x years of age. When scaling the main and interaction effects PGSs by division with $s_{main} := \frac{1}{k}\sum_{l=1}^{k} |\hat{\beta}_{G_l}^{(int)}|$ or $s_{Gxage} := \frac{1}{k}\sum_{l=1}^{k} |\hat{\beta}_{G_l*age}^{(int)}|$, respectively, the sum of the two PGSs results in

$$(\sum_{j=1}^{k} \frac{|\hat{\beta}_{G_j}^{(int)}|}{s_{main}} * G_j) + (\sum_{j=1}^{k} \frac{|\hat{\beta}_{G_j*age}^{(int)}|}{s_{G_jxage}} * G_j) * (x-50)$$

This compares to

$$\sum_{j=1}^{k}(\frac{|\hat{\beta}_{G_j}^{(int)}|}{(s_{main} + s_{Gxage})} + \frac{|\hat{\beta}_{G_j*age}^{(int)}|}{(s_{main} + s_{Gxage})}(x-50) * G_j)$$

which is the age-informative PGS using scaled weights under the above stated assumptions. When testing the age-informative PGS in a linear model (*model 2*),

$$Y \sim \gamma_{PGS} * (\sum_{j=1}^{k}(\frac{|\hat{\beta}_{G_j}^{(int)}|}{(s_{main} + s_{Gxage})} + \frac{|\hat{\beta}_{G_j*age}^{(int)}|}{(s_{main} + s_{Gxage})}(x-50) * G_j))$$

this is comparable to the *Jayasinghe* et al.[1] model (*model 3*),

$$Y \sim \gamma_{PGS}^{main} * (\sum_{j=1}^{k} \frac{|\hat{\beta}_{G_j}^{(int)}|}{s_{main}} * G_j) + \gamma_{PGS*age}^{Gxage} * (\sum_{j=1}^{k} \frac{|\hat{\beta}_{G_j*age}^{(int)}|}{s_{Gxage}} * G_j) * AGE50$$

except *model 3* obtains two separate $\gamma$-estimates for main effect PGS and interaction effect PGS[1].

**Supplemental Figure S1. Differences and similarities of distribution for age-agnostic and age-informative PGS across 282 kidney function SNPs in UK Biobank and KORA&AugUR.** We derived age-agnostic and age-informative PGSs based on 282 kidney function-related SNPs. Shown are density plots of unscaled and scaled PGSs stratified by age groups in (**a**) UK Biobank and (**b**) KORA&AugUR.

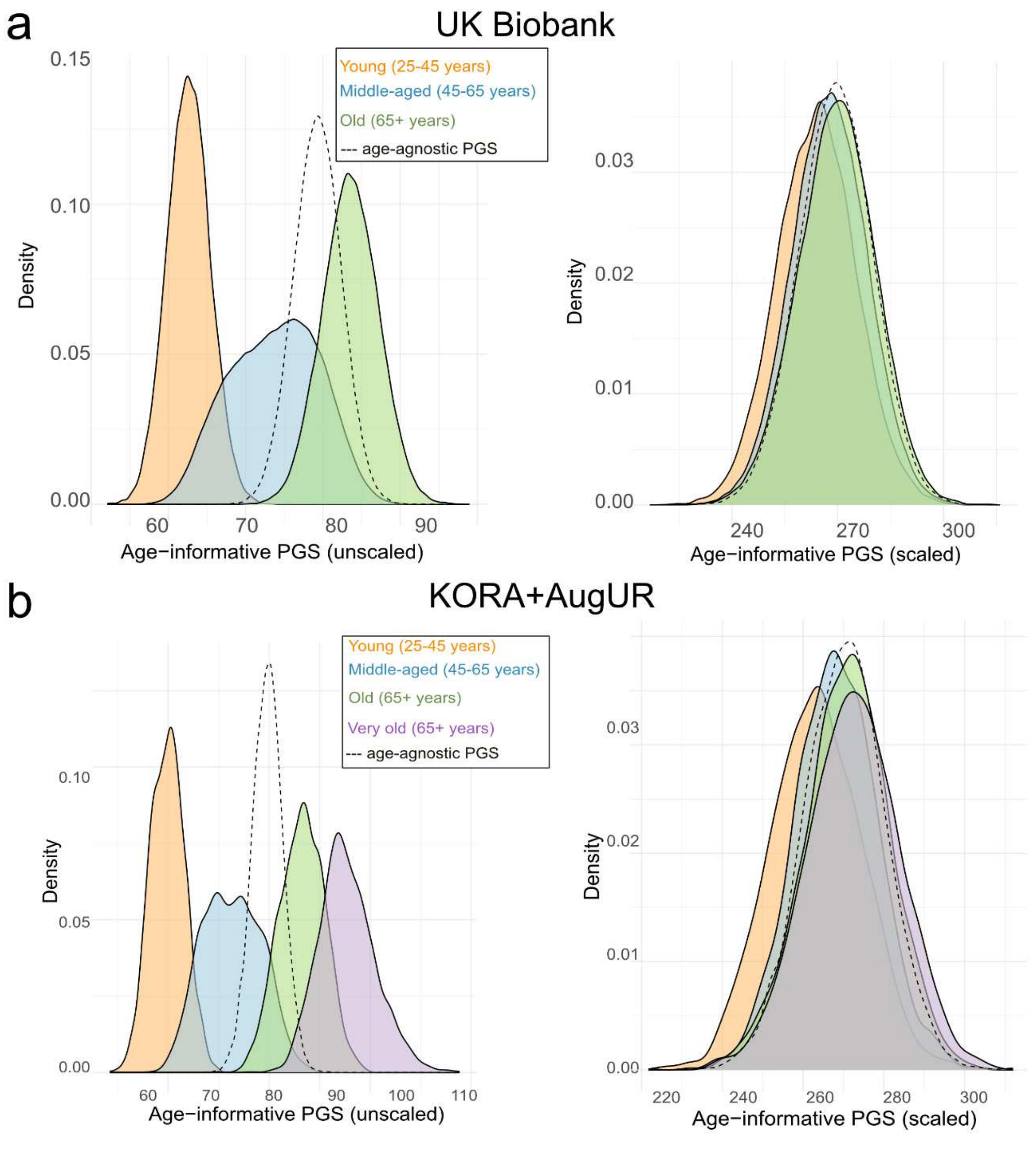

### Supplemental Tables

**Supplemental Table S1. Weights for unscaled and scaled PGS in the theoretical example**. In a simplified case study, we considered five SNPs without interaction on trait Y (SNP 1-5, light grey) and five SNPs with age interaction (SNP 6-10, dark grey) where genetic effects start at age 50. For the age-independent SNPs, we set $\hat{\beta}_{G_j}^{(int)}$ = -0.5 trait units per allele and thus $\hat{\beta}_{G_j}^{(age)}$= -0.5 trait units per allele for any age. For the age-dependent SNPs, we set $\hat{\beta}_{G_j}^{(age)}$ to 0 until the age of 50 years and $\hat{\beta}_{G_j xage}^{(int)}$= -0.05 trait units per allele and year and thus $\hat{\beta}_{Gj}^{(age=x)} = 0 - 0.05 * (x - 50) for\ x \geq 50$. The effect allele is defined as the Y-lowering allele assuming that this is the "bad" allele. Unscaled weights are derived from beta-estimates as $w_j = |\hat{\beta}_{Gj}|$ or $w_j^{(age=x)} = |\hat{\beta}_{Gj}^{(int)} + \hat{\beta}_{Gj*Age}^{(int)} * (x - 50)$ | or scaled by the average of these weights. The variance of the age-agnostic PGS is derived as $(\sum_{j=1}^{k} w_j{}^2) * var(G)$, while for age-informative PGS at a given age (x years), it is $(\sum_{j=1}^{k} w_j^{(age=x)^2}) * var(G)$ assuming constant variance of SNPs and constant variance of Y across age (var(Y)=130). For the scaled PGS, the $R^2$ is derived as $\gamma_{PGS}^2 * \frac{var(PGS_k^{age})}{var(Y)}$, where the average of scaled weights reflects the per PGS unit effect on trait Y ($\gamma_{PGS}$).

| | $w_j$ | $w_j^{(age=40)}$ | $w_j^{(age=50)}$ | $w_j^{(age=60)}$ | $w_j^{(age=70)}$ | $w_j^{(age=80)}$ |
|---|---|---|---|---|---|---|
| **Unscaled weights** | | | | | | |
| Age-independent ("stable") SNPs | | | | | | |
| SNP1 | 0.5 | 0.5 | 0.5 | 0.5 | 0.5 | 0.5 |
| SNP2 | 0.5 | 0.5 | 0.5 | 0.5 | 0.5 | 0.5 |
| SNP3 | 0.5 | 0.5 | 0.5 | 0.5 | 0.5 | 0.5 |
| SNP4 | 0.5 | 0.5 | 0.5 | 0.5 | 0.5 | 0.5 |
| SNP5 | 0.5 | 0.5 | 0.5 | 0.5 | 0.5 | 0.5 |
| Age-dependent ("decline") SNPs | | | | | | |
| SNP6 | 0.6 | 0 | 0 | 0.5 | 1 | 1.5 |
| SNP7 | 0.6 | 0 | 0 | 0.5 | 1 | 1.5 |
| SNP8 | 0.6 | 0 | 0 | 0.5 | 1 | 1.5 |
| SNP9 | 0.6 | 0 | 0 | 0.5 | 1 | 1.5 |
| SNP10 | 0.6 | 0 | 0 | 0.5 | 1 | 1.5 |
| **Average weight** | 0.55 | 0.25 | 0.25 | 0.5 | 0.75 | 1 |
| **Var(PGS)** | 1.525 | 0.625 | 0.625 | 1.25 | 3.125 | 6.25 |
| **Scaled weights** | | | | | | |
| SNP1 | 0.9 | 2 | 2 | 1 | 0.7 | 0.5 |
| SNP2 | 0.9 | 2 | 2 | 1 | 0.7 | 0.5 |
| SNP3 | 0.9 | 2 | 2 | 1 | 0.7 | 0.5 |
| SNP4 | 0.9 | 2 | 2 | 1 | 0.7 | 0.5 |
| SNP5 | 0.9 | 2 | 2 | 1 | 0.7 | 0.5 |
| SNP6 | 1 | 0 | 0 | 1 | 1.3 | 1.5 |
| SNP7 | 1 | 0 | 0 | 1 | 1.3 | 1.5 |
| SNP8 | 1 | 0 | 0 | 1 | 1.3 | 1.5 |
| SNP9 | 1 | 0 | 0 | 1 | 1.3 | 1.5 |
| SNP10 | 1 | 0 | 0 | 1 | 1.3 | 1.5 |
| **Average weight** | 1 | 1 | 1 | 1 | 1 | 1 |
| **Var($G_j$)=Var(G)** | 0.5 | 0.5 | 0.5 | 0.5 | 0.5 | 0.5 |
| **Var(PGS)** | 5 | 10 | 10 | 5 | 5.5 | 6.8 |
| **Var(Y)** | 130 | 130 | 130 | 130 | 130 | 130 |
| **$R^2$(PGS)** | 3.8% | 7.7% | 7.7% | 3.8% | 4.2% | 5.2% |

**Supplemental Table S2. Age-specific beta-estimates for 12 age-dependent SNPs in UK Biobank.** We derived age-specific beta-estimates for the 12 known SNPs with significant age-interaction on eGFR ($P_{Gj*age}$ <0.05/595=8.4x10$^{-5}$). Shown are published marginal ($\hat{\beta}_{Gj}$), main ($\hat{\beta}_{Gj}^{(int)}$) and interaction effects ($\hat{\beta}_{Gj*age}^{(int)}$) for each SNP[2]. For age = x, we derived the age-specific beta-estimate $\hat{\beta}_{Gj}^{(age=x)} = \hat{\beta}_{Gj}^{(int)} + \hat{\beta}_{Gj*age}^{(int)}(x-50)$. Beta-estimates correspond to the effect allele (EA) defined as the marginal eGFR-lowering allele (i.e. $\hat{\beta}_{Gj}$ <0). We show the age when $\hat{\beta}_{Gj}^{(age)} = 0$ as "starting age" of the genetic effect. The unit of beta-estimates is mL/min/1.73m² per allele or mL/min/1.73m² per allele and per year for the interaction effects.

| SNP | Locus name | EA | $\hat{\beta}_{G_j}$ | $\hat{\beta}_{G_j}^{(int)}$ | $\hat{\beta}_{G_j*age}^{(int)}$ | $\hat{\beta}_{G_j}^{(40)}$ | $\hat{\beta}_{G_j}^{(50)}$ | $\hat{\beta}_{G_j}^{(60)}$ | $\hat{\beta}_{G_j}^{(70)}$ | $\hat{\beta}_{G_j}^{(80)}$ | Starting Age (years) |
|---|---|---|---|---|---|---|---|---|---|---|---|
| **SNPs with age-dependent effect (decline SNPs)** | | | | | | | | | | | |
| rs77924615 | *UMOD/PDILT* | G | -0.905 | -0.382 | -0.06 | -0.442 | -0.985 | | -1.588 | -2.191 | 43.7 |
| rs13334589 | *UMOD/PDILT* | A | -0.832 | -0.376 | -0.054 | -0.43 | -0.919 | | -1.461 | -2.003 | 43.1 |
| rs10224002 | *PRKAG2* | G | -0.71 | -0.601 | -0.014 | -0.615 | -0.741 | | -0.882 | -1.023 | 7.3 |
| rs74209810 | *UMOD/PDILT* | T | -0.427 | -0.216 | -0.032 | -0.248 | -0.536 | | -0.856 | -1.176 | 43.3 |
| rs2076668 | *GGT7* | A | -0.379 | -0.255 | -0.014 | -0.269 | -0.391 | | -0.526 | -0.661 | 31.1 |
| rs434215 | *TPPP* | A | -0.372 | -0.166 | -0.026 | -0.192 | -0.426 | | -0.686 | -0.946 | 43.6 |
| rs4930319 | *OVOL1* | C | -0.288 | -0.174 | -0.014 | -0.188 | -0.314 | | -0.454 | -0.594 | 37.6 |
| rs854922 | *RRAGD* | A | -0.283 | -0.06 | -0.024 | -0.084 | -0.303 | | -0.547 | -0.791 | 47.6 |
| rs2783971 | *SDCCAG8* | A | -0.238 | -0.109 | -0.018 | -0.127 | -0.285 | | -0.46 | -0.635 | 43.8 |
| rs1458038 | *FGF5* | C | -0.238 | -0.127 | -0.014 | -0.141 | -0.263 | | -0.399 | -0.534 | 40.6 |
| rs2921093 | *PRAG1* | T | -0.222 | -0.124 | -0.012 | -0.136 | -0.248 | | -0.371 | -0.495 | 40.0 |
| rs28857283 | *C15orf54* | G | -0.127 | 0.032 | -0.019 | 0.014 | -0.154 | | -0.34 | -0.526 | 48.3 |
| **Average effect** | | | -0.418 | -0.213 | -0.025 | -0.238 | -0.464 | | -0.714 | -0.965 | NA |

**Supplemental Table S3. Descriptive statistics of the age-agnostic and age-informative PGS by study and age group.** We generated age-agnostic and age-informative PGSs in UK Biobank and KORA&AugUR using unscaled and scaled weights. Shown are mean values and standard deviation (SD) of PGSs in the overall sample and by age group.

| Age range (years) | **Overall** | **Young** 25-45 | **Middle-aged** 45-65 | **Old** 65+* | **Very old** 75-95 |
|---|---|---|---|---|---|
| **UK Biobank, n** | 254,068 | 41,099 | 276,096 | 115,103 | - |
| Age-agnostic PGS | | | | | |
| unscaled | 79.3 (3.1) | 79.3 (3.1) | 79.3 (3.1) | 79.3 (3.1) | - |
| scaled | 269.4 (10.5) | 269.4 (10.5) | 269.4 (10.5) | 269.4 (10.5) | - |
| Age-informative PGS | | | | | |
| unscaled | 74.7 (7.6) | 62.3 (2.9) | 73.2 (5.9) | 85.5 (4.6) | - |
| scaled | 267.6 (10.9) | 263.9 (11.2) | 267.5 (10.7) | 270.2 (11.1) | - |
| **KORA&AugUR, n** | 9,001 | 2,079 | 4,012 | 3,283 | 2,807 |
| Age-agnostic PGS | | | | | |
| unscaled | 79.7 (3.0) | 79.7 (3.0) | 79.7 (3.0) | 79.7 (3.0) | 79.7 (3.0) |
| scaled | 270.3 (10.1) | 270.3 (10.1) | 270.3 (10.1) | 270.3 (10.1) | 270.3 (10.1) |
| Age-informative PGS | | | | | |
| unscaled | 77.8 (13.9) | 59.9 (3.3) | 71.7 (5.8) | 86.4 (4.2) | 95.8 (5.5) |
| scaled | 268.5 (11.4) | 263.2 (11.3) | 267.9 (10.3) | 270.9 (10.5) | 272.6 (11.2) |

*KORA&AugUR: 65-75 years; n= number of individuals.

**Supplemental Table S4. Association and predictive ability of age-agnostic and age-informative PGS for kidney function in cross-sectional data**. We derived the age-agnostic and age-informative PGSs based on 282 kidney function-related SNPs in UK Biobank and KORA&AugUR. We derived the PGS association of the age-agnostic and the age-informative PGS via linear regression (*model 1:* $eGFR \sim \gamma_0 + \gamma_{PGS} * PGS_{282} + \gamma_{age} * AGE50 + \varepsilon$ and *model 2:* $eGFR \sim \gamma_0 + \gamma_{PGS} * PGS_{282}^{age} + \gamma_{age} * AGE50 + \varepsilon$ , respectively). We contrasted results with a model including only AGE50 (*model 0*). Shown are the variance of eGFR, the residual variance of eGFR adjusted for age, effect estimates with 95%-CIs and p-values. Also shown is the partial $R^2$ of the PGS derived as $R^2_{total} - \frac{R^2_{reduced}}{1-R^2_{reduced}}$ with $R^2_{reduced}$ obtained from *model 0* and total $R^2$ of the model. Effect estimates (γ) correspond to the effect on eGFR per year (age) or per allele with average effect on eGFR (PGS). The effect allele is the eGFR-lowering allele of the marginal effect.

| | | Age agnostic | Age-informative |
|---|---|---|---|
| **Model name** | **Model 0** | **Model 1** | **Model 2** |
| *Covariables* | *AGE50* | *AGE50, $PGS_{282}$* | *AGE50, $PGS_{282}^{age}$* |
| **UK Biobank (n=254,068)** | | | |
| **eGFR** | | | |
| Var(eGFR) | 163.9 | 163.9 | 163.9 |
| Var(eGFR\|age) | 130.5 | 130.5 | 130.5 |
| Intercept | 99.6 | 167.7 | 165.7 |
| **Age** | | | |
| γ (95%-CI) | -0.71 (-0.72; -0.71) | -0.71 (-0.72; -0.71 | -0.72 (-0.73; -0.72) |
| P | <2e-16 | <2e-16 | <2e-16 |
| **$PGS_{282}$** | | | |
| γ (95%-CI) | - | -0.25 (-0.26 -0.25) | -0.25 (-0.25; -0.24) |
| P | - | <2e-16 | <2e-16 |
| $R^2$ | - | **5.4%** | **5.5%** |
| **Total $R^2$** | 20.4% | 24.7% | 24.7% |
| **KORA&AugUR (n=9,001)** | | | |
| **eGFR** | | | |
| Var(eGFR) | 370.8 | 370.8 | 370.8 |
| Var(eGFR\|age) | 196.2 | 196.2 | 196.2 |
| Intercept | 95.9 | 168.2 | 163.0 |
| **Age** | | | |
| γ (95%-CI) | -0.79 (-0.81; -0.77) | -0.79 (-0.81; -0.78) | -0.73 (-0.75; -0.72) |
| P | <2e-16 | <2e-16 | <2e-16 |
| **$PGS_{282}$** | | | |
| γ (95%-CI) | - | -0.27 (-0.30; -0.24) | -0.25 (-0.28; -0.23) |
| P | - | <2e-16 | <2e-16 |
| $R^2$ | - | **3.7%** | **3.9%** |
| **Total $R^2$** | 47.1% | 49.0% | 49.2% |

Age was centered at 50 years.

**Supplemental Table S5. Association and predictive ability of age-agnostic and age-informative PGS for kidney function by age group in cross-sectional data**. We derived the age-agnostic and age-informative PGSs based on 282 kidney function-related SNPs in UK Biobank and KORA&AugUR. We derived the PGS association of the age-agnostic and the age-informative PGS by age groups via linear regression (*model 1:* $eGFR \sim \gamma_0 + \gamma_{PGS} * PGS_{282} + \gamma_{age} * AGE50 + \varepsilon$ and *model 2:* $eGFR \sim \gamma_0 + \gamma_{PGS} * PGS_{282}^{age} + \gamma_{age} * AGE50 + \varepsilon$ , respectively). We contrasted results with a model including only AGE50 (*model 0*). Shown are the variance of eGFR, the residual variance of eGFR adjusted for age, effect estimates with 95%-CIs and p-values. Also shown is the partial $R^2$ of the PGS derived as $R^2_{total} - \frac{R^2_{reduced}}{1-R^2_{reduced}}$ with $R^2_{reduced}$ obtained from *model 0* and total $R^2$ of the model. Effect estimates (γ) correspond to the effect on eGFR per year (age) or per allele with average effect on eGFR (PGS). The effect allele is the eGFR-lowering allele of the marginal effect.

| | | Age agnostic | Age-informative |
|---|---|---|---|
| **Model name** | **Model 0** | **Model 1** | **Model 2** |
| *Covariables* | *AGE50* | *AGE50, $PGS_{282}$* | *AGE50, $PGS_{282}^{age}$* |
| **UK Biobank** | | | |
| **Young (25-45 years), n=41,099** | | | |
| **eGFR** | | | |
| Var(eGFR) | 125.9 | 125.9 | 125.9 |
| Var(eGFR\|age) | 123.6 | 123.6 | 123.6 |
| Intercept | 99.6 | 158.0 | 154.6 |
| **Age** | | | |
| γ (95%-CI) | -0.67 (-0.72; -0.62) | -0.67 (-0.72; -0.62) | -0.69 (-0.75; -0.65) |
| P | <2e-16 | <2e-16 | <2e-16 |
| **$PGS_{282}$** | - | | |
| γ (95%-CI) | - | -0.22 (-0.22; -0.21) | -0.21 (-0.22; -0.20) |
| P | - | <2e-16 | <2e-16 |
| $R^2$ | - | **4.2%** | **4.4%** |
| **Total $R^2$** | 1.8% | 5.9% | 6.1% |
| **Middle-aged (45-65 years), n=276,096** | | | |
| **eGFR** | | | |
| Var(eGFR) | 135.9 | 135.9 | 135.9 |
| Var(eGFR\|age) | 131.1 | 131.1 | 131.1 |
| Intercept | 99.1 | 163.1 | 162.0 |
| **Age** | | | |
| γ (95%-CI) | -0.67 (-0.68; -0.66) | -0.67 (-0.68; -0.66) | -0.67 (-0.68; -0.66) |
| P | <2e-16 | <2e-16 | <2e-16 |
| **$PGS_{282}$** | - | | |
| γ (95%-CI) | - | -0.24 (-0.24; -0.23) | -0.23 (-0.24; -0.23) |
| P | - | <2e-16 | <2e-16 |
| $R^2$ | - | **4.9%** | **5.0%** |
| **Total $R^2$** | 11% | 15.2% | 15.3% |
| **Old-aged (65-80 years), n= 115,103** | | | |
| **eGFR** | | | |
| Var(eGFR) | 189.3 | 189.3 | 189.3 |
| Var(eGFR\|age) | 177.0 | 177.0 | 177.0 |
| Intercept | 104.1 | 176.7 | 173.9 |
| **Age** | | | |
| γ (95%-CI) | -1.07 (-1.1; -1.05) | -1.07 (-1.1; -1.05) | -1.08 (-1.10; -1.06) |
| P | <2e-16 | <2e-16 | <2e-16 |
| **$PGS_{282}$** | - | | |
| γ (95%-CI) | - | -0.27 (-0.28; -0.26) | -0.25 (-0.26; -0.25) |
| P | - | <2e-16 | <2e-16 |
| $R^2$ | - | **4.5%** | **4.7%** |
| **Total $R^2$** | 6.5% | 10.7% | 10.9% |

| *Table continued* | **Model 0**<br>*AGE50* | **Model 1**<br>*AGE50, $PGS_{282}$* | **Model 2**<br>*AGE50, $PGS_{282}^{age}$* |
|---|---|---|---|
| **KORA&AugUR** | | | |
| **Young (25-45 years), n=2,079** | | | |
| **eGFR** | | | |
| Var(eGFR) | 174.1 | 174.1 | 174.1 |
| Var(eGFR\|age) | 163.9 | 163.9 | 163.9 |
| Intercept | 97.1 | 180.4 | 173.3 |
| **Age** | | | |
| γ (95%-CI) | -0.61 (-0.72; -0.51) | -0.61 (-0.71; -0.50) | -0.62 (-0.72; -0.52) |
| P | <2e-16 | <2e-16 | <2e-16 |
| **$PGS_{282}$** | - | | |
| γ (95%-CI) | - | -0.31 (-0.36; -0.25) | -0.30 (-0.35; -0.25) |
| P | - | <2e-16 | <2e-16 |
| $R^2$ | - | **5.9%** | **6.3%** |
| **Total $R^2$** | 5.8% | 11.3% | 11.6% |
| **Middle-aged (45-65 years), n=4,012** | | | |
| **eGFR** | | | |
| Var(eGFR) | 177.3 | 177.3 | 177.3 |
| Var(eGFR\|age) | 160.8 | 160.8 | 160.8 |
| Intercept | 96.3 | 169.0 | 166.7 |
| **Age** | | | |
| γ (95%-CI) | -0.70 (-0.76; -0.63) | -0.70 (-0.77; -0.63) | -0.71 (-0.77; -0.64) |
| P | <2e-16 | <2e-16 | <2e-16 |
| **$PGS_{282}$** | - | | |
| γ (95%-CI) | - | -0.27 (-0.31; -0.23) | -0.26 (-0.30; -0.23) |
| P | - | <2e-16 | <2e-16 |
| $R^2$ | - | **4.6%** | **4.5%** |
| **Total $R^2$** | 9.3% | 13.5% | 13.4% |
| **Old-aged (65-75 years), n= 3,283** | | | |
| **eGFR** | | | |
| Var(eGFR) | 211.8 | 211.8 | 211.8 |
| Var(eGFR\|age) | 205.8 | 205.8 | 205.8 |
| Intercept | 97.3 | 173.9 | 170.5 |
| **Age** | | | |
| γ (95%-CI) | -0.85 (-1.01; -0.68) | -0.85 (-1.02; -0.69) | -0.86 (-1.03; -0.69) |
| P | <2e-16 | <2e-16 | <2e-16 |
| **$PGS_{282}$** | | | |
| γ (95%-CI) | - | -0.28 (-0.33; -0.23) | -0.26 (-0.30; -0.22) |
| P | - | <2e-16 | <2e-16 |
| $R^2$ | - | **3.7%** | **3.9%** |
| **Total $R^2$** | 2.8% | 6.4% | 6.5 % |
| **Very-old-aged (75-98 years), n= 2,807** | | | |
| **eGFR** | | | |
| Var(eGFR) | 291.6 | 291.6 | 291.6 |
| Var(eGFR\|age) | 277.4 | 277.4 | 277.4 |
| Intercept | 97.5 | 158.7 | 159.3 |
| **Age** | | | |
| γ (95%-CI) | -0.91 (-1.06; -0.76) | -0.90 (-1.05; -0.75) | -0.92 (-1.07; -0.78) |
| P | <2e-16 | <2e-16 | <2e-16 |
| **$PGS_{282}$** | | | |
| γ (95%-CI) | - | -0.23 (-0.29; -0.17) | -0.22 (-0.27; -0.16) |
| P | - | <2e-16 | <2e-16 |
| $R^2$ | - | **1.8%** | **2.3%** |
| **Total $R^2$** | 4.8% | 6.6% | 7.0% |

Age was centered at 50 years.

**Supplemental Table S6. Association and predictive ability of main and interaction-effect PGS for kidney function in *Jayasinghe* models**. We derived the main effect PGS and the interaction effect PGS, $PGS_{282}^{(main)}$ and $PGS_{282}^{(Gxage)}$,based on 282 kidney function-related SNPs in UK Biobank and KORA&AugUR. We derived the PGS association of the main effects PGS and the interaction effects PGS via linear regression (*model 3:* $eGFR \sim \gamma_0 + \gamma_{PGS}^{main} * PGS_{282}^{main} + \gamma_{PGS*age}^{Gxage} * PGS_{282}^{Gxage} * AGE50 + \gamma_{age} * AGE50 + \varepsilon$ and *model 4:* $eGFR \sim \gamma_0 + \gamma_{PGS} * PGS_{282}^{age} + \gamma_{age} * AGE50 + \gamma_{PGS}^{Gxage} * PGS_{282}^{Gxage} + \varepsilon$). We compared the results to a model including the age-informative PGS (*model 2*, **Supplemental Table 4**). Shown are the variance of eGFR, the residual variance of eGFR adjusted for age, effect estimates with 95%-CIs and p-values. Also shown is the partial $R^2$ of the PGS derived as $R_{total}^2 - \frac{R_{reduced}^2}{1-R_{reduced}^2}$ with $R_{reduced}^2$ obtained from *model 0* (**Supplemental Table 4**) and total $R^2$ of the model. Thus, partial $R^2$ provides the predictive ability of $PGS_{282}^{main}$, $PGS_{282}^{Gxage}$ (*model 3*) and additionally *AGE50*, $PGS_{282}^{Gxage}$ (*model 4*). Effect estimates (γ) correspond to the effect on eGFR per year (age) or per allele with average effect on eGFR (PGS). The effect allele is the eGFR-lowering allele of the marginal effect.

| | Jayasinghe approach | |
|---|---|---|
| **Model name**<br>*Covariables* | **Model 3**<br>$PGS_{282}^{main}$, $PGS_{282}^{Gxage} * AGE50$ | **Model 4**<br>$PGS_{282}^{main}$, $PGS_{282}^{Gxage} * AGE50$, $PGS_{282}^{Gxage}$ |
| **UK Biobank (n=254,068)** | | |
| Intercept | 163.3 | 163.2 |
| **Main effect** (Age) | | |
| γ (95%-CI) | -0.14 (-0.18; -0.10) | -0.17 (-0.23; -0.12) |
| P | $3.7x10^{-10}$ | $<2x10^{-16}$ |
| **Main effect** ($PGS_{282}^{main}$) | | |
| γ (95%-CI) | -0.24 (-0.24; -0.23) | -0.24 (-0.24; -0.24) |
| P | $<2x10^{-16}$ | $<2x10^{-16}$ |
| **Main effect** ($PGS_{282}^{Gxage}$) | | |
| γ | - | -0.0019( -0.041; -0.00016) |
| P | - | $<2x10^{-16}$ |
| **Interaction effect** ($PGS_{282}^{Gxage}$xAge) | | |
| γ (95%-CI) | -0.0020 (-0.0022; -0.0018) | -0.0019 (-0.0021; -0.0017) |
| P | $<2x10^{-16}$ | $<2x10^{-16}$ |
| $R^2$ (PGS) | **5.5%** | **5.5%** |
| Total $R^2$ | 24.8% | 24.8% |
| **KORA&AugUR (n=9,001)** | | |
| Intercept | 161.1 | 163.9 |
| **Main effect** (Age) | | |
| γ (95%-CI) | -0.25 (-0.42; -0.08) | -0.37 (-0.56; -0.18) |
| P | 0.0032 | $1.2x10^{-4}$ |
| **Main effect** ($PGS_{282}^{(main)}$) | | |
| γ | -0.24 (-0.27; -0.22) | -0.24 (-0.26; -0.21) |
| P | $<2x10^{-16}$ | $<2x10^{-16}$ |
| **Main effect** ($PGS_{282}^{Gxage}$) | | |
| γ | - | -0.0017( -0.029; -0.00044) |
| P | - | $8.5x10^{-3}$ |
| **Interaction effect** ($PGS_{282}^{Gxage}$xAge) | | |
| γ (95%-CI) | -0.0019 (-0.0025; -0.0013) | -0.0015 (-0.0021; -0.00080) |
| P | $1.5x10^{-10}$ | $1.3x10^{-5}$ |
| $R^2$ (PGS) | **3.9%** | **4.0%** |
| Total $R^2$ | 49.1% | 49.2% |

Age was centered at 50 years.

**Supplemental Table S7. Performance of main and interaction-effects PGS for kidney function in *Jayasinghe* models**. We derived the age-informative, the main effect PGS and the interaction effect PGS, as well as the age-agnostic PGS based on 282 kidney function-related SNPs in UK Biobank and KORA&AugUR. We derived the PGS association of the main effects PGS and the interaction effects PGS via linear mixed models according to the *Jayasinghe* approach[1] (*model 3L:* $eGFR_t \sim \gamma_0 + RI + \gamma_{PGS}^{main} * PGS_{282}^{main} + \gamma_{PGS*age}^{Gxage} * PGS_{282}^{Gxage} * AGE50_t + \gamma_{age} * AGE50_t + RS + \varepsilon$ and *model 4L:* $eGFR_t \sim \gamma_0 + RI + \gamma_{PGS}^{main} * PGS_{282}^{main} + \gamma_{PGS*age}^{Gxage} * PGS_{282}^{Gxage} * AGE50_t + \gamma_{age} * AGE50_t + RS + \gamma_{PGS}^{Gxage} * PGS_{282}^{Gxage} + \varepsilon$). We derived the association of the age-agnostic and the age-informative PGS adding their interaction with age (*extended model 1L:* $eGFR_t \sim \gamma_0 + RI + \gamma_{PGS} * PGS_{282} + \gamma_{PGS*age} * PGS_{282} * AGE50_t + \gamma_{age} * AGE50_t + RS + \varepsilon$ and *extended model 2L:* $eGFR_t \sim \gamma_0 + RI + \gamma_{PGS} * PGS_{282}^{age} + \gamma_{PGS*age} * PGS_{282}^{age} * AGE50_t + \gamma_{age} * AGE50_t + RS + \varepsilon$, respectively). Shown are the variance of eGFR, the residual variance of eGFR adjusted for age, effect estimates with 95%-CIs and p-values. Also shown is the partial (marginal) $R^2$ of the PGS derived as $R_{total}^2 - \frac{R_{reduced}^2}{1-R_{reduced}^2}$ with $R_{reduced}^2$ obtained from a model including only AGE50 and total (marginal) $R^2$ of the model. Thus, partial $R^2$ provides the predictive ability of $PGS_{282}^{main}$, $PGS_{282}^{Gxage}$ (*model 3L*) and additionally *AGE50*, $PGS_{282}^{Gxage}$ (*model 4L*). Effect estimates (γ) correspond to the effect on eGFR per year (age) or per allele with average effect on eGFR (PGS). The effect allele is the eGFR-lowering allele of the marginal effect.

| | Age-informative | Jayasinghe approach | | Age-agnostic |
|---|---|---|---|---|
| **Model name**<br>*Covariables* | **Model 2L extended**<br>*Age50, $PGS_{282}^{age}$, $PGS_{282}^{age}$* AGE50* | **Model 3L**<br>*$PGS_{282}^{(main)}$, $PGS_{282}^{(Gxage)}$* AGE50* | **Model 4L**<br>*$PGS_{282}^{(main)}$, $PGS_{282}^{(Gxage)}$* AGE50, $PGS_{282}^{(Gxage)}$* | **Model 1L extended**<br>*AGE50, $PGS_{282}$, $PGS_{282}$* AGE50* |
| **UK Biobank (n=348,275, m=1,520,382)** | | | | |
| Intercept | 151.2 | 161.0 | 161.2 | 161.3 |
| **Main effect** (Age) | | | | |
| γ (95%-CI) | 0.80 (0.70;0.90) | -0.24 (-0.24; -0.23) | -0.20 (-0.24;-0.15) | -0.14 (-0.24;-0.03) |
| P | <2x10$^{-16}$ | <2x10$^{-16}$ | <2x10$^{-16}$ | 0.012 |
| **Main effect** (*$PGS_{282}^{age}$*/$PGS_{282}^{main}$/ $PGS_{282}$) | | | | |
| γ (95%-CI) | -0.19 (-0.19;-0.18) | -0.24 (-0.23; -0.22) | -0.23 (-0.24;-0.23) | -0. 22 (-023;-0. 22) |
| P | <2x10$^{-16}$ | <2x10$^{-16}$ | <2x10$^{-16}$ | <2x10$^{-16}$ |
| **Main effect** ($PGS_{282}^{(Gxage)}$) | | | | |
| γ (95%-CI) | - | - | 0. 0028 (0.001;0.004) | - |
| P | - | - | 0.002 | - |
| **Interaction effect** ($PGS_{282}^{(Gxage)}$xAge) | | | | |
| γ (95%-CI) | -0.0061 (-0.0065;-0.0057) | -0.0022 (-0.0024;-0.0020) | -0.0024 (-0.0025; -0.0022) | -0.0028 (-0.0031; -0.0023) |
| P | 2.6x10$^{-5}$ | <2x10$^{-16}$ | <2x10$^{-16}$ | <2x10$^{-16}$ |
| $R^2$ (PGS) | **4.7%** | **4.3%** | **4.3%** | **4.1%** |
| Total $R^2$ | - | 27.0% | 27.0% | 26.8% |
| **KORA&AugUR (n=9,057, m=16,804)** | | | | |
| Intercept | 158.2 | 161.1 | 165.3 | 171.2 |
| **Main effect** (Age) | | | | |
| γ (95%-CI) | -0.18 (-0.55;-0.19) | -0.28 (-0.43; -0.19) | -0.40 (-0.57; -0.23) | -1.02 (-1.44; -0.60) |
| P | 0.32 | 3.2x10$^{-4}$ | 6.8x10$^{-6}$ | 2x10$^{-6}$ |
| **Main effect** (*$PGS_{282}^{age}$*/$PGS_{282}^{main}$/ $PGS_{282}$) | | | | |
| γ (95%-CI) | -0.25 (-0.27,-0.23) | -0.24 (-0.27;-0.22) | -0.24 (-0.27;-0.21) | -0. 28 (-0. 31;-0. 25) |
| P | <2x10$^{-16}$ | <2x10$^{-16}$ | <2x10$^{-16}$ | <2x10$^{-16}$ |
| **Main effect** ($PGS_{282}^{Gxage}$) | | | | |
| γ (95%-CI) | - | - | -0. 16 (-0.29; -0.056) | - |
| P | - | - | 3.5x10$^{-3}$ | - |
| **Interaction effect** ($PGS_{282}^{Gxage}$xAge) | | | | |
| γ (95%-CI) | -0.0022 (-0.0035;-0.00087) | -0.0018 (-0.00025; -0.00013) | -0.0014 (-0.0024; -0.0013) | 0.00078 (-0.00080;0.0023) |
| P | 0.0015 | 1.5x10$^{-10}$ | 3.5x10$^{-6}$ | 0.33 |
| $R^2$ (PGS) | **3.7%** | **3.9%** | **3.9%** | **3.6%** |
| Total $R^2$ | 45.9% | 49.1% | 46.1% | 46.0% |

Age was centered at 50 years.

**Supplemental Table S8. Predictive ability of age-agnostic and interaction-effect PGS for eGFR-decline using person-specific slopes in a two-step approach.** We derived the age-agnostic PGS and the interaction effect PGS based on the 282 kidney function SNPs in UK Biobank and KORA&AugUR. We applied a two-step approach to analyze the PGS association with eGFR-decline: first, we estimated person-specific slopes via "best linear unbiased predictors" (BLUPs) using a linear mixed model with random intercepts and random slopes, $eGFR_t \sim AGE50_t$ in longitudinal data. Second, we tested the association of each PGS with person-specific slopes (PSS) as the outcome via univariable linear regression in cross-sectional data ($PSS \sim \gamma_0 + \gamma_{PGS} * PGS_{282} + \varepsilon$ or $PSS \sim \gamma_0 + \gamma_{PGS} * PGS_{282}^{Gxage} + \varepsilon$, respectively). Shown are PGS effect estimates with respective 95% CIs in mL/min/1.73m² per allele per year. Also shown is the marginal R2 of the model.

| | **Age-agnostic PGS** $PGS_{282}$ | **Interaction-effect PGS** $PGS_{282}^{(Gxage)}$ |
|---|---|---|
| **UK Biobank (n=254,068)** | | |
| Intercept | -0.55 (-0.58; -0.52) | -0.75 (-0.76; -0.74) |
| **PGS** | | |
| γ (95%CI) | -0.0012 (-0.13; -0.11) | -0.00044 (-0.00048; -0.000041) |
| P | <2e-16 | <2e-16 |
| $R^2$ | **0.2%** | **0.3%** |
| **KORA&AugUR (n=9,001)** | | |
| Intercept | -0.38 (-0.51; -0.25) | -0.59 (-0.65; -0.54) |
| **PGS** | | |
| γ (95%CI) | -0.0016 (-0.0021; -0.0011) | -0.00075 (-0.00094; -0.00057) |
| P | $1 \times 10^{-10}$ | $2.4 \times 10^{-14}$ |
| $R^2$ | **0.7%** | **1%** |

Age was centered at 50 years.

**Supplemental Table S9. Association of genetic and non-genetic risk factors with change of eGFR over age.** We stratified genetic risk groups by quintiles of the interaction effect PGS (Q1-Q5) in KORA&AugUR. We tested the association of these quintiles and their interaction with age in longitudinal data. We applied a linear mixed model without (*model 3Q*: $eGFR_t \sim \gamma_0 + RI + \gamma_{PGS}^{main} * PGS_{282}^{main} + \sum_{j=2}^{5} \gamma_j * I_{Q_j^{Gxage}} * AGE50_t + \gamma_{age} * AGE50_t + RS + \varepsilon$) and with the inclusion of non-genetic risk factors and their interaction with age (*model 3Q extended*). $I_{Q_j^{Gxage}}$ is the indicator variable of the PGS quintile. Shown are effect estimates in mL/min/1.73m² (main effects) or mL/min/1.73m² per year (interaction effect) with 95%-CIs. The intercept provides mean eGFR for the reference group and the age effect is the mean annual decline of the reference group (50-year-old person with lowest genetic risk (Q1) or 50-year-old person with normal weight, no diabetes nor albuminuria, and lowest genetic risk (Q1), respectively). The main effect of a risk factor can be interpreted as the change of eGFR-level when this risk factor is present. The interaction effect of risk factor with age is the annual decline for individuals with this risk factor additional to the annual decline in the reference group. Marginal $R^2$ for fixed effects is given as total $R^2$. marginal $R^2$ for fixed effects is given as total $R^2$.

| | **Model 3Q** | **Model 3Q extended** |
|---|---|---|
| n/m | 9,057/16,804 | 7,988/10,742 |
| Intercept | 162.6 | 98.6 |
| **Main effects** | | |
| Age | -0.75 (-0.77; -0.71) | -0.66 (-0.73; -0.64) |
| $PGS_{282}^{main}$ | -0.24 (-0.28; -0.22) | -0.23 (-0.26; -0.20) |
| Diabetes | - | 1.68 (-0.32; 3.68) |
| Obesity | - | -1.92 (-2.87; -0.99) |
| Overweight | - | -1.81 (-2.55; -1.07) |
| Albuminuria | - | -1.93 (-3.34; -0.52) |
| **Interaction effects** | | |
| Age x Diabetes | - | -0.23 (-0.31; -0.15) |
| Age x Overweight | - | -0.18 (-0.06; 0.035) |
| Age x Obesity | - | -0.12 (-0.17; -0.069) |
| Age x Albuminuria | - | -0.039 (-0.098;0.020) |
| Age x Q2 | -0.040 (-0.083, 0.0038) | -0.056 (-0.11; -0.007) |
| Age x Q3 | -0.088 (-0.13; -0.044) | -0.10 (-0.15; -0.050) |
| Age x Q4 | -0.09 (-0.13; -0.047) | -0.11 (-0.16; -0.061) |
| Age x Q5 | -0.13 (-0.18; -0.09) | -0.15 (-0.17; -0.047) |
| Total $R^2$ | 44.6% | 46.5% |

Diabetes was defined via self-report, antidiabetic medication, or HbA1c ≥6.5%. Albuminuria was defined as UACR ≥30 mg/g. BMI>25 kg/m² and <30 kg/m² was defined as “overweight” and ≥30kg/m² as “obese”. n=number of individuals; m=number of measurements. Age was centered at 50 years.

### Supplemental References

1. Jayasinghe D, Momin MM, Beckmann K, Hyppönen E, Benyamin B, Lee SH. Mitigating type I error inflation and power loss in GxE PRS: Genotype-environment interaction in polygenic risk score models. Genet Epidemiol. 2024;48:85–100. doi:10.1002/gepi.22546.

2. Wiegrebe S, Gorski M, Herold JM, Stark KJ, Thorand B, Gieger C, et al. Analyzing longitudinal trait trajectories using GWAS identifies genetic variants for kidney function decline. Nat Commun. 2024;15:10061. doi:10.1038/s41467-024-54483-9.
